# EEG Correlates of Delayed Emergence after Remimazolam-Induced Anaesthesia Compared to Propofol

**DOI:** 10.1101/2024.05.17.24307522

**Authors:** Yeji Lee, Sujung Park, Hyoungkyu Kim, Jeongmin Kim, Eun Jung Kim, Youngjai Park, Uncheol Lee, Jeongwook Kwon, Joon-Young Moon, Bon-Nyeo Koo

**Affiliations:** Center for Neuroscience Imaging Research, Institute for Basic Science (IBS), Suwon 16419, Republic of Korea; Sungkyunkwan University (SKKU), Suwon 16419, Republic of Korea; Department of Anaesthesiology and Pain Medicine, Yonsei University College of Medicine; Anaesthesia and Pain Research Institute; Research institute of Slowave Inc., Seoul, South Korea; Department of Anaesthesiology and Pain Medicine, Asan Medical Center, University of Ulsan College of Medicine, Seoul, Republic of Korea; Center for Consciousness Science, Department of Anaesthesiology, University of Michigan Medical School, Ann Arbor, USA

**Author notes:** co-1^st^ (Yeji Lee, Sujung Park, Hyoungkyu Kim).

**Keywords:** delayed emergence prediction, EEG phase analysis, frontal EEG, propofol TIVA, recovery of cognition, remimazolam TIVA

## Abstract

**Background:** Delayed emergence from anaesthesia presents clinical challenges, including prolonged stays in the post-anaesthesia care unit (PACU). The neurobiological mechanisms underlying delayed emergence, particularly in remimazolam-induced anaesthesia, remain poorly understood. This study aimed to elucidate these mechanisms by comparing remimazolam and propofol anaesthesia, focusing on prefrontal electroencephalogram (EEG).

**Methods:** Patients (age > 18, *n* = 48) underwent laparoscopic cholecystectomy randomly received remimazolam or propofol general anaesthesia. Power spectrogram analysis and functional connectivity measures, phase lag entropy (PLE) and phase lag index (PLI), were employed to the prefrontal EEG data collected at baseline, unconsciousness, and emergence. Correlation between EEG measures and Patient State Index (PSI) at PACU, as well as time to Aldrete 9, were compared.

**Results:** PSI values (*P* < 0.0001, *P* = 0.006) and time to Aldrete 9 at PACU (*P* < 0.001) revealed slower recovery in remimazolam-induced anaesthesia. Remimazolam group exhibited residual effects in power at theta (*P* = 0.018) and alpha (*Ps* < 0.001) bands and lower PLE during emergence in the alpha (*P* < 0.0001, *P* = 0.015) and beta (*P* = 0.016, *P* < 0.001) bands. Delayed consciousness recovery (time to Aldrete 9) under remimazolam was significantly correlated with PLE (Pearson’s *r* = -.78, *P* < 0.0001), and PLI (Pearson’s *r* = .69, *P* = 0.028) in the alpha band during deep anaesthesia.

**Conclusion:** Dynamic changes in prefrontal EEG and the correlation analyses show the potential of EEG in predicting emergence speed, providing insights into the neurobiological mechanisms of short-term delayed emergence in remimazolam anaesthesia.

## Introduction

Delayed emergence from anaesthesia is a situation in which a patient may take longer than expected to regain consciousness, movement, and spontaneous breathing. This can lead to extended stays at the post-anaesthesia care unit (PACU), increased healthcare resource utilization, decreased patient satisfaction, and, in rare cases, require careful differential diagnosis with serious medical problems^1^.

The neurobiological mechanisms underlying delayed emergence are related to neurotransmitter imbalances, neuroinflammation, and altered brain connectivity^1^. Delayed emergence may also be a sign of an underlying medical condition or intraoperative event. It is therefore essential to carefully manage the anaesthetic to minimize the risk. Comparative studies of remimazolam versus propofol anaesthesia have reported delayed emergence after remimazolam administration^2–4^. Remimazolam, a newly introduced ultra-short-acting benzodiazepine sedative, acts through GABA receptors in a manner similar to propofol^5^. However, the neurobiological mechanisms are poorly understood due to the lack of comprehensive studies of EEG oscillation patterns during emergence.

The difference in emergence time between propofol and remimazolam is likely due to dose and the presence or absence of flumazenil^2, 4, 6–8^. Flumazenil can rapidly reverse remimazolam sedation, potentially surpassing propofol in recovery speed^9^. Studies have recommended lower remimazolam doses for certain patient populations to mitigate the risk of delayed emergence^3^.

Under anaesthesia, the levels of consciousness are associated with decreased variability in inter-regional communication in the brain^10–12^. Changes in functional connectivity and changes in EEG power, particularly in prefrontal regions, are used as indicators of level of consciousness^11, 12^. Exploring the dynamic changes in EEG activity during emergence from general anaesthesia is essential to gain valuable insights into the brain states that transition from unconsciousness to consciousness^13^. GABA receptor-specific anaesthetics, such as propofol, produce a variety of EEG patterns while anaesthesia is maintained. These include an increase in prefrontal alpha EEG power, a shift in EEG power to lower frequencies, and changes in coherence^14–18^. Despite the importance of EEG studies, there is a lack of knowledge about the changes in EEG characteristics and functional connectivity during remimazolam-induced anaesthesia.

This study aimed to elucidate the mechanisms underlying the delayed emergence from remimazolam. We compared remimazolam and propofol anaesthesia, focusing on prefrontal EEG oscillation patterns during baseline, unconsciousness, and emergence. Our results showed significant between-group differences in power by frequency band, functional connectivity, and PSI values. We also revealed associations between functional connectivity and cognitive recovery, which means that the time to cognitive recovery can be predicted in remimazolam-induced anaesthesia.

## Methods

### Patient population

This prospective, randomized, double-blinded study was performed between June 2023 and October 2023. The study protocol was approved by the institutional review board of Severance Hospital, Yonsei University Health System. After obtaining ethics committee approval, all patients were informed written consents on the day before surgery and the protocol was done in accordance with relevant guidelines.

The inclusion criteria were as follows: Patients aged 19 to 65 years who meet the American Society of Anaesthesiologists (ASA) physical status classification 1-3 and undergo laparoscopic cholecystectomy. The exclusion criteria were as follows: Pregnant women, individuals with arrhythmias, emergency surgeries, obesity (BMI > 30), day surgery, foreigners, and illiterate individuals.

### Study procedure

Among the 60 patients enrolled in this study, individuals with low PSI scores at baseline, insufficient EEG data, or a lack of marker flag were excluded. Consequently, we analysed the data from the remaining 48 patients (n = 24 per group) assigned patients to the remimazolam or propofol group at a 1:1 ratio (Figure 1A). Patients, operators, and two research personnel were unaware of the group identity, but the attending anaesthesiologist couldn’t be blinded about the group identity due to the distinct properties and color of the two anaesthetics.

**Figure 1.**
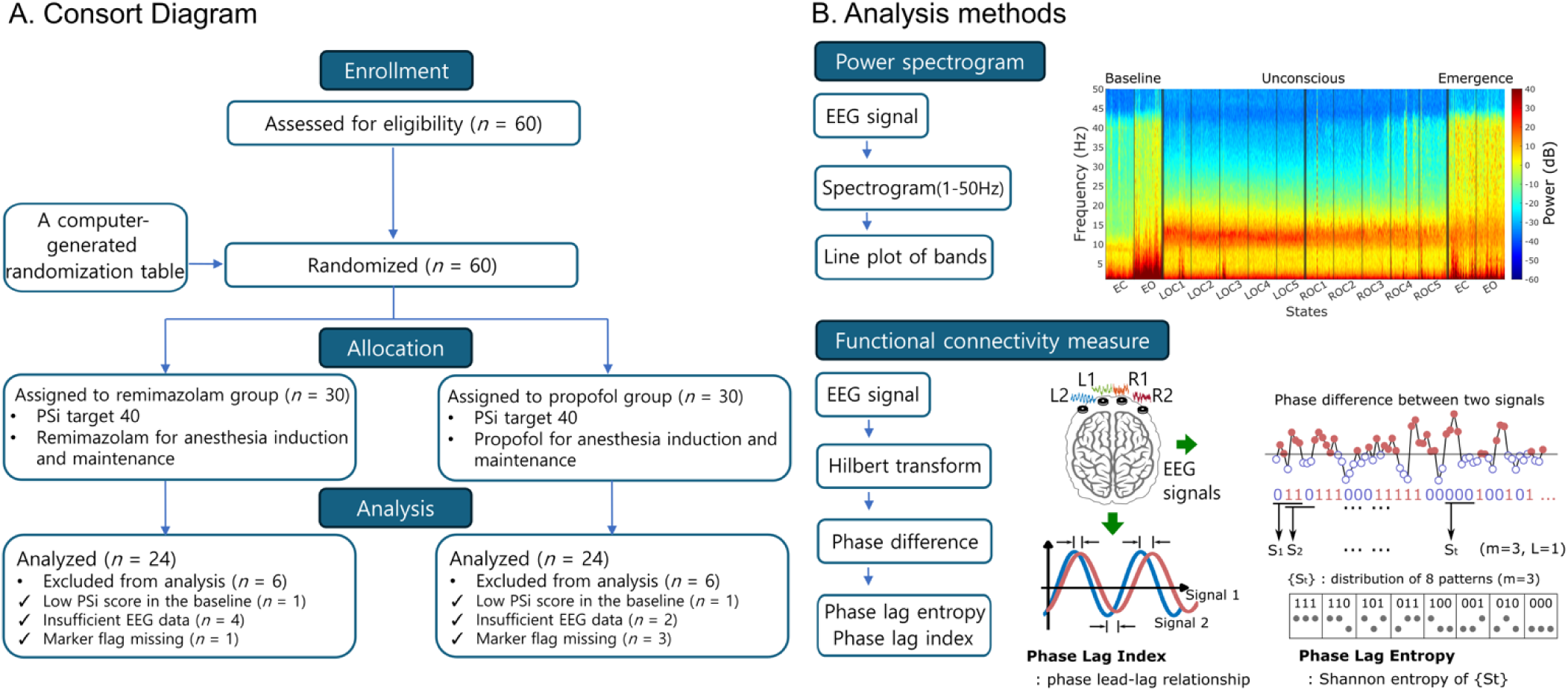
Schematic diagram. (A) Consort diagram describes the progress of patient via each stage of the randomized trial. (B) Analysis methods describe the progress of the analysis methods which include power spectrogram analysis and functional connectivity measure analysis.

Before entering the operating rooms, we placed the SEDLine sensor on the patient’s forehead. Preoperative baseline EEG were recorded at controlled eye-open (focused wakefulness) and eye-closed (relaxed wakefulness) conditions, for each 4-minute period.

Upon entering the operating rooms, the patients were monitored with pulse oximetry, non-invasive blood pressure, electrocardiography, and anaesthetic depth (SedLine^®^, Masimo Corp., Irvine, CA, USA). The patients received 0.1 mg glycopyrrolate before infusing remifentanil and remimazolam or propofol using a commercial syringe pump (Agillia, SB Medica SRL, Italy).

In the propofol group, anaesthesia was induced using propofol [target-controlled infusion (TCI), Marsh model] and remifentanil at effect-site concentrations of 4 mcg ml^-1^ and 3 ng ml^-1^ respectively. The concentration of propofol is adjusted to maintain an appropriate depth of anaesthesia based on the EEG (PSI target 40) until the end of the surgery. In the remimazolam group, anaesthesia was induced using remifentanil at an effect-site concentration of 3 ng ml^-1^ (TCI, Minto model) and remimazolam at a flow rate of 6 mg kg^-1^ h^-1^, as per the manufacturer’s recommendations. The concentration of remimazolam is adjusted to maintain an appropriate depth of anaesthesia based on the EEG (PSI target 40) until the end of the surgery. More details are provided in Supplementary Method (‘Study Procedure’).

### EEG Data pre-processing

A four-channel (Fp1(L1), Fp2(R1), F7(L2), F8(R2)) EEG signals (*n* = 60; 30 in each group) were acquired during the entire anaesthetic process, including baseline, unconscious, and emergence periods. As EEG characteristics are distinctly exhibited in different anaesthetic states, the data were divided into 6 states. EEGLAB open-source toolbox for MATLAB^19^ was used for data extraction by states. (1) baseline eyes-closed (**EC**): defined as 3 minutes after eyes-closed; (2) baseline eyes-open (**EO**): defined as 3 minutes after eyes-open; (3) after loss of consciousness (**after LOC**): defined as 15 minutes after the loss of verbal command response following the administration of propofol or remimazolam; (4) before recovery of consciousness (**before ROC**): defined as 15 minutes before the extubation point; (5) eyes-closed (**EC**): defined as 3 minutes after patients closed their eyes at PACU; (6) eyes-open (**EO**): defined as 3 minutes after patients opened their eyes at the PACU. To compare the prefrontal EEG characteristics of two different anaesthetic agents under the unconsciousness period in stages, the periods after LOC and before ROC were divided into five intervals, aligning with the duration of baseline EO and EC states for significance assessments.

The EEG data of each state was segmented into a 10-sec time series (with a uniform epoch length), which generated a total of 18 epochs (3*60)/10 for each subject and each band.

### Prefrontal EEG data analyses

The prefrontal EEG data was analysed using MATLAB and band-pass filtered within the range of 1-50Hz. The analysis included the power spectrogram which was focused on five frequency bands (delta: 0.5-4 Hz, theta: 4-8 Hz, alpha: 8-13 Hz, beta: 13-20 Hz, gamma: 20-40 Hz). Details of power spectrogram analysis and timeseries power analysis are provided in the Supplementary Method (‘Power Spectrogram and Line Power Analyses’). The second section provided in the Supplementary Method (‘Functional Connectivity Measure Analysis’) includes the functional connecitivity analyses (Phase lag entropy and phase lag index) and evaluates the association between the functional connectivity and the clinical scores from propofol- and remimazolam-induced anaesthesia.

### Statistical analysis

To evaluate the group differences between propofol-and remimazolam-induced anaesthesia, statistical analyses were performed in RStudio (R version 4.3.2). A two-way repeated-measures analysis of variance was conducted to compare the PSI, power, and functional connectivity between the two groups at 14 different states. Post-hoc analysis with Bonferroni correction was applied to adjust for multiple comparisons. For all data analyses, the corrected *P*-values of less than 0.05 were deemed to represent a statistically significant difference between the two groups (†*P* < 0.0001; \*\*\**P* < 0.001; \*\**P* < 0.01; \**P* < 0.05). Pearson correlation coefficients (r) were calculated using the MATLAB (version R2022b, Mathworks Inc., Co., Ltd., USA) to see the correlation between EEG measures (i.e., connectivity and powers at four frequency bands) and indices values (i.e., PSI scores at PACU (averaged score of EC and EO emergence) and time to Aldrete 9). Benjamini-Hochberg (FDR) correction for multiple testing was applied for the correlations of all comparisons.

## Results

### Study procedure, participant overview and analytical methodology

Our analysis methods included power spectrogram analysis across various frequency bands, such as delta (0.5-4 Hz), theta (4-8 Hz), alpha (8-13 Hz), beta (13-20 Hz), and gamma (20-40 Hz). Functional connectivity measures were incorporated into the analysis method. We utilized phase lag entropy (PLE) to assess the entropy of relative phase patterns and the phase lag index (PLI) to evaluate phase lead-lag relationships (Figure 1B).

The demographic data of the 24 participants in each group showed no significant differences between the groups, and similar results were observed for the use of remifentanil, operative time, anaesthetic time, blood pressure 5 minutes after intubation, heart rate, and blood pressure 5 minutes before extubation (Table 1). However, patients administered remimazolam exhibited evident sedation and reduced consciousness level at the PACU, reflected by lower Aldrete scores and a RASS, and required a longer duration to meet PACU discharge criteria compared to those receiving propofol (Table 1).

**Table 1.**
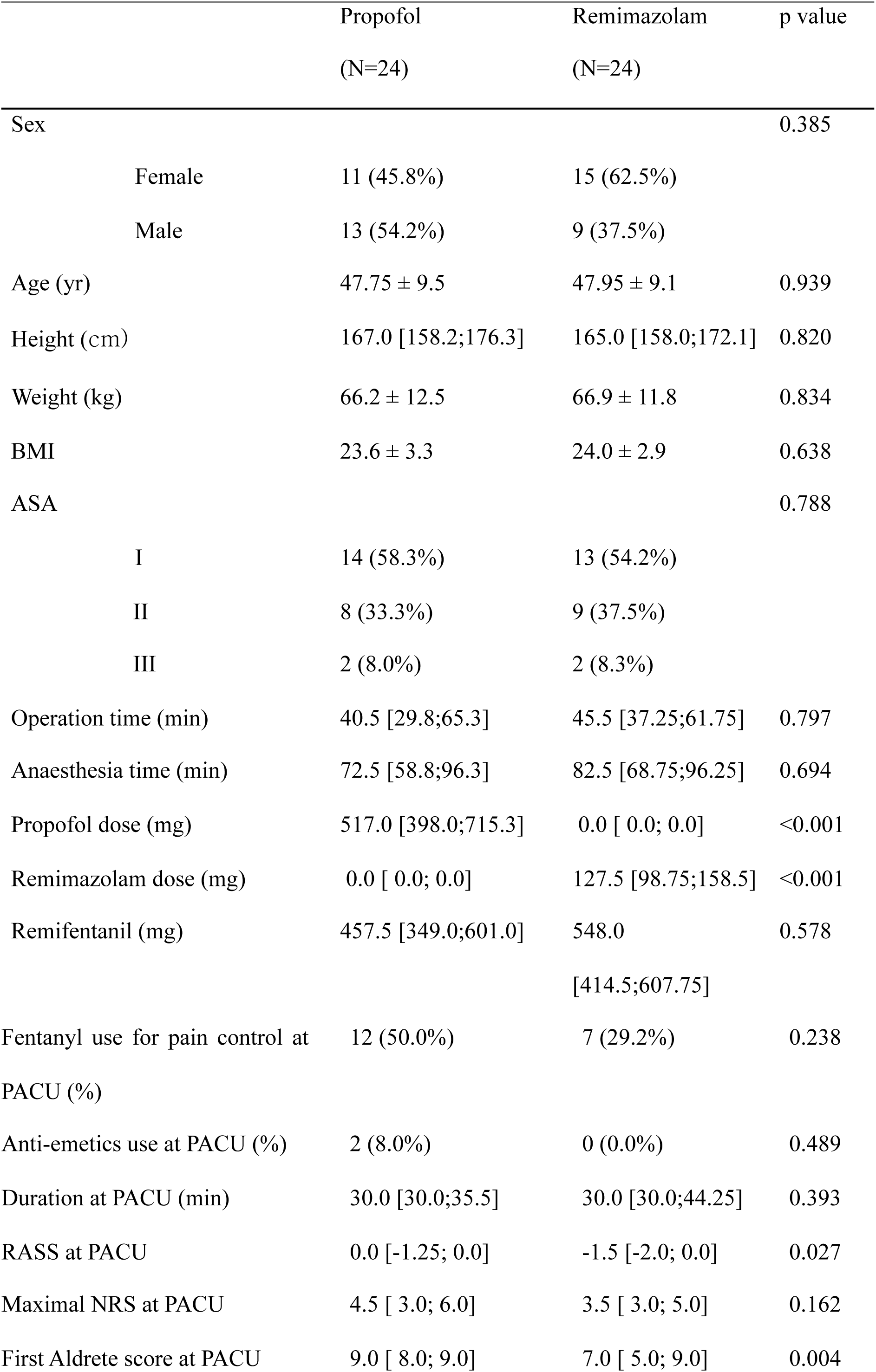
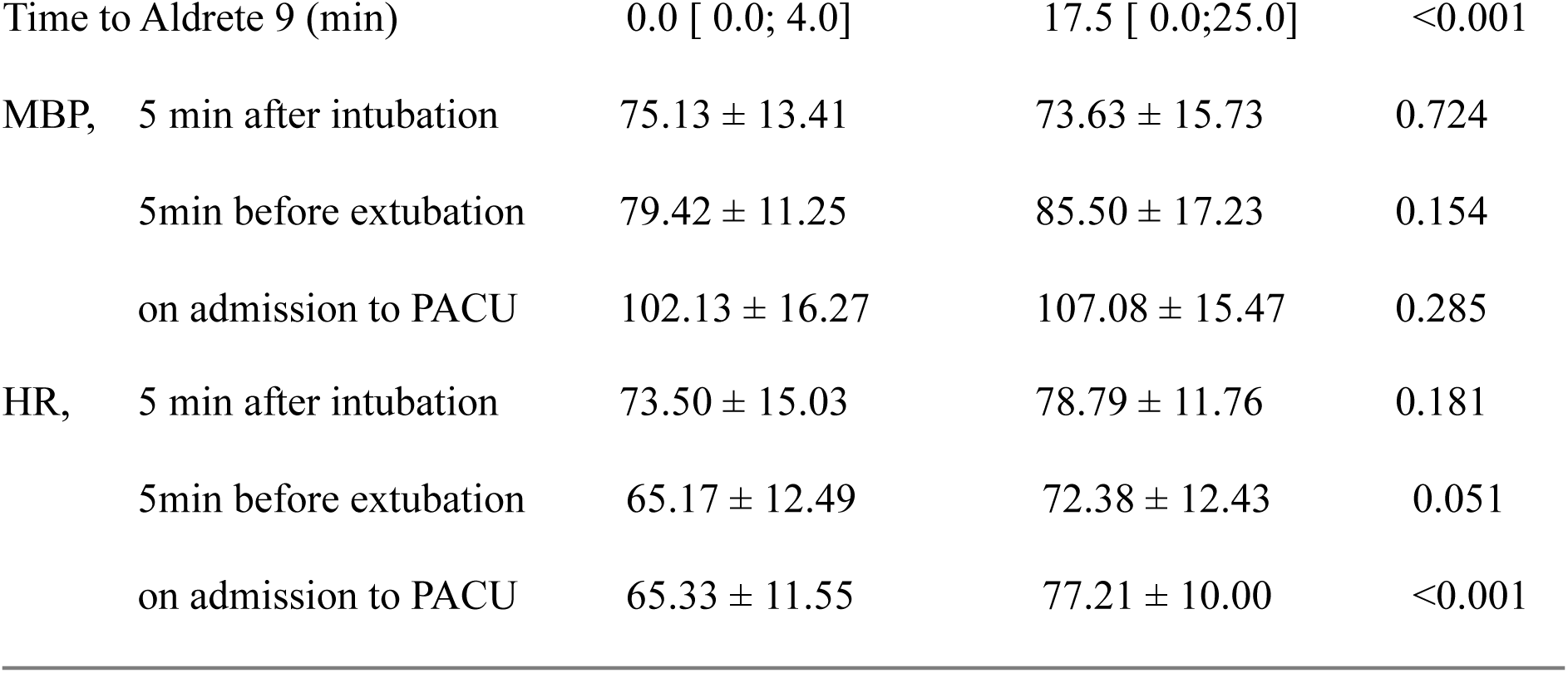
Patients’ demographics and characteristics of intraoperative and post-operative information. Values indicate number (%), means (±SD), or median [inter-quartile range]. BMI: body mass index; ASA: American Society of Anaesthesiologists. RASS scores evaluate alertness (0 = alert, negative value = drowsy); Aldrete score evaluate movement, breathing, and vital sign (+9 = fully recovered); Time to Aldrete 9 indicates the time taken for full emergence. PACU: post-anaesthesia care unit; RASS: Richmond Agitation-Sedation Scale; NRS: numerical rating scale for pain.

|  | Propofol<br>(N=24) | Remimazolam<br>(N=24) | p value |
| --- | --- | --- | --- |
| Sex |  |  | 0.385 |
| Female | 11 (45.8%) | 15 (62.5%) |  |
| Male | 13 (54.2%) | 9 (37.5%) |  |
| Age (yr) | 47.75 ± 9.5 | 47.95 ± 9.1 | 0.939 |
| Height (cm) | 167.0 [158.2;176.3] | 165.0 [158.0;172.1] | 0.820 |
| Weight (kg) | 66.2 ± 12.5 | 66.9 ± 11.8 | 0.834 |
| BMI | 23.6 ± 3.3 | 24.0 ± 2.9 | 0.638 |
| ASA |  |  | 0.788 |
| I | 14 (58.3%) | 13 (54.2%) |  |
| II | 8 (33.3%) | 9 (37.5%) |  |
| III | 2 (8.0%) | 2 (8.3%) |  |
| Operation time (min) | 40.5 [29.8;65.3] | 45.5 [37.25;61.75] | 0.797 |
| Anaesthesia time (min) | 72.5 [58.8;96.3] | 82.5 [68.75;96.25] | 0.694 |
| Propofol dose (mg) | 517.0 [398.0;715.3] | 0.0 [ 0.0; 0.0] | <0.001 |
| Remimazolam dose (mg) | 0.0 [ 0.0; 0.0] | 127.5 [98.75;158.5] | <0.001 |
| Remifentanyl (mg) | 457.5 [349.0;601.0] | 548.0<br>[414.5;607.75] | 0.578 |
| Fentanyl use for pain control at PACU (%) | 12 (50.0%) | 7 (29.2%) | 0.238 |
| Anti-emetics use at PACU (%) | 2 (8.0%) | 0 (0.0%) | 0.489 |
| Duration at PACU (min) | 30.0 [30.0;35.5] | 30.0 [30.0;44.25] | 0.393 |
| RASS at PACU | 0.0 [-1.25; 0.0] | -1.5 [-2.0; 0.0] | 0.027 |
| Maximal NRS at PACU | 4.5 [ 3.0; 6.0] | 3.5 [ 3.0; 5.0] | 0.162 |
| First Aldrete score at PACU | 9.0 [ 8.0; 9.0] | 7.0 [ 5.0; 9.0] | 0.004 |
| Time to Aldrete 9 (min) | 0.0 [ 0.0; 4.0] | 17.5 [ 0.0;25.0] | <0.001 |
| MBP, 5 min after intubation | 75.13 ± 13.41 | 73.63 ± 15.73 | 0.724 |
| 5min before extubation | 79.42 ± 11.25 | 85.50 ± 17.23 | 0.154 |
| on admission to PACU | 102.13 ± 16.27 | 107.08 ± 15.47 | 0.285 |
| HR, 5 min after intubation | 73.50 ± 15.03 | 78.79 ± 11.76 | 0.181 |
| 5min before extubation | 65.17 ± 12.49 | 72.38 ± 12.43 | 0.051 |
| on admission to PACU | 65.33 ± 11.55 | 77.21 ± 10.00 | <0.001 |

### Power spectrogram analysis

In our power spectrogram analysis, remimazolam-induced anaesthesia (Figure 2B) exhibited residual effects (alpha power from 10Hz to 13 Hz) compared to propofol-induced anaesthesia (Figure 2A) during the EO and EC periods of emergence. This power differential was further confirmed in the power (dB) ratio between the two anaesthetic agents (Figure 2C). Around alpha band (8-13Hz), higher propofol power was evident before regaining consciousness (ROC), and higher remimazolam power was noted during emergence (EC, EO) period. Within the beta band (13-20Hz), higher power was consistently observed throughout the unconscious and emergence periods in the remimazolam group. In the gamma band (20-40 Hz), higher power was observed during ROC4-ROC5, as well as during EC and EO emergence in the propofol group. Time-series plots of powers at lower frequency bands exhibited significant differences in anaesthetic drug effects across the anaesthesia process. Specifically, the delta band (Figure 2D) indicated significantly higher power in the propofol group relative to the remimazolam group in the ROC1 (*P* = 0.039). Theta power (Figure 2E) of propofol was significantly higher throughout the before-ROC period (*P* = 0.006, *P* = 0.013, *P* = 0.004, *P* < 0.001, *P* = 0.002), with the opposite observed during the EO emergence period (*P* = 0.018). Alpha power (Figure 2F) revealed significantly lower levels in the propofol group compared to the remimazolam group during the EC and EO emergence periods (*Ps* < 0.001). However, no anaesthetic drug difference was observed at the beta and gamma power (Figure 2G, 2H).

**Figure 2.**
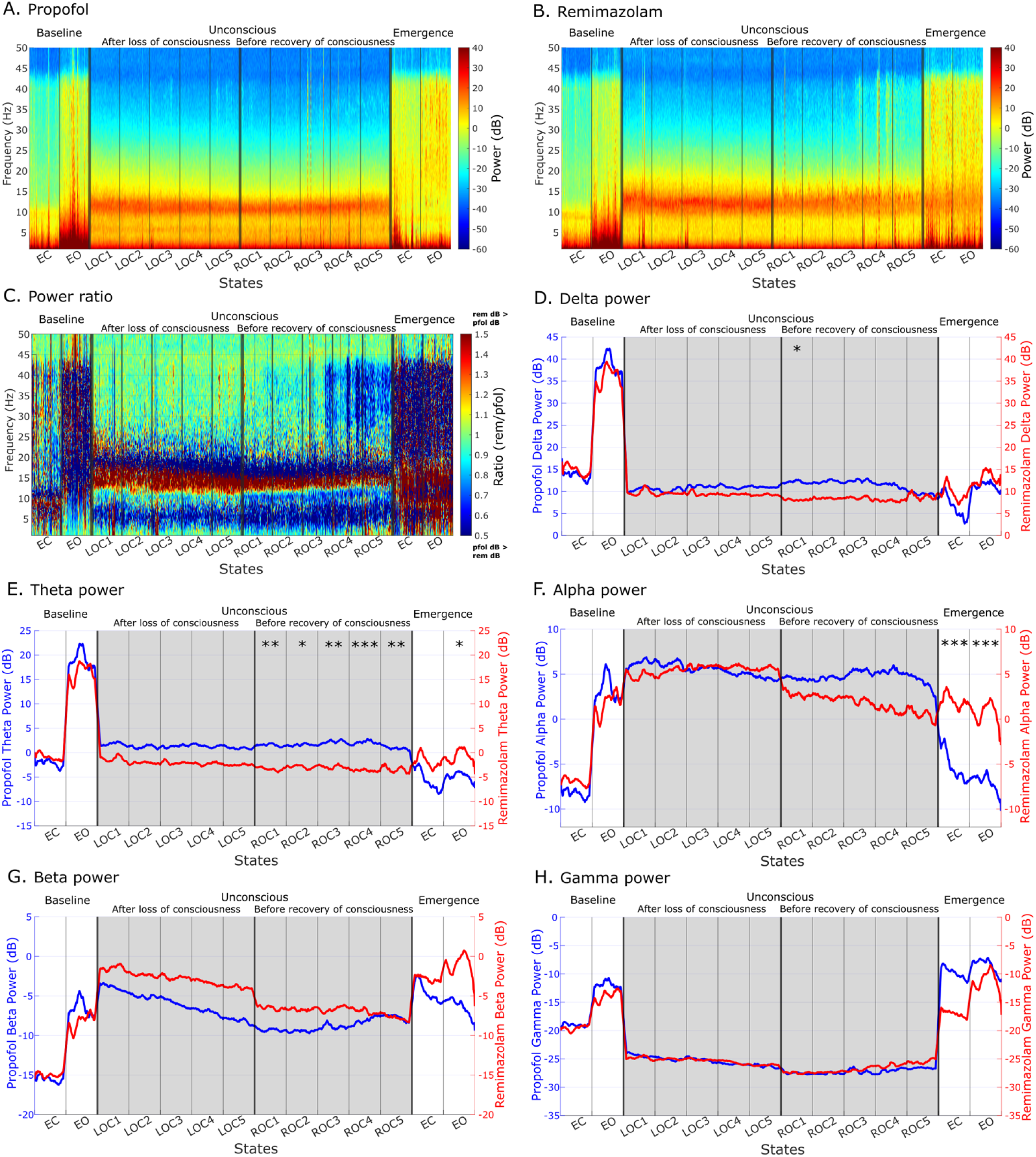
Power comparison between propofol (n=24) and remimazolam (n=24) on the prefrontal region (F8). Averaged power spectrograms of (A) propofol-induced anaesthesia and (B) remimazolam-induced anaesthesia at 14 states. (C) Power ratio of two anaesthetic drugs. Time-series plots of powers across the different anaesthesia periods at (D) delta, (E) theta, (F) alpha, (G) beta, and (H) gamma frequency bands. Group and time interaction showed significant effects under all five frequency band powers. †*P* < 0.0001, \*\*\**P* < 0.001, \*\**P* < 0.01, \**P* < 0.05 in post-hoc analysis.

### Functional connectivity analysis

The results of functional connectivity analyses elucidated distinct prefrontal EEG features between the remimazolam and propofol groups across different frequency bands (Figure 3). PLE, the diversity of phase lead/lag patterns, progressively increased from baseline through the unconscious period in the theta band (4-8 Hz) but decreased in the alpha and beta bands (8-20 Hz). During the unconscious period, the drug difference in PLE was notable across frequency bands, with remimazolam anaesthesia exhibiting higher PLE in the theta and alpha bands, but the opposite finding in the beta band. Specifically, differences were found throughout the after-LOC period (*P* = 0.006, *P* = 0.02, *P* < 0.001, *P* < 0.001, *P* < 0.0001) and ROC1-ROC4 (*P* < 0.001, *P* = 0.008, *P* = 0.008, *P* = 0.036) from the unconscious period in the theta band (Figure 3A); ROC1-ROC4 (*P* = 0.007, *P* < 0.001, *P* < 0.001, *P* = 0.001) from the unconscious period in the alpha band (Figure 3C); LOC1 (*P* = 0.015), LOC3-LOC5 (*P* = 0.008, *P* = 0.001, *P* < 0.0001) and ROC1-ROC3 (*P* = 0.001, *P* < 0.001, *P* = 0.017) from the unconscious period in the beta band (Figure 3E). We identified residual anteriorization and delayed arousal during emergence following remimazolam-induced anaesthesia compared to propofol anaesthesia, leading to decreased entropy of phase lead/lag patterns (alpha: *P* < 0.0001, *P* = 0.015; beta: *P* = 0.016, *P* < 0.001; Figure 3C & 3E).

**Figure 3.**
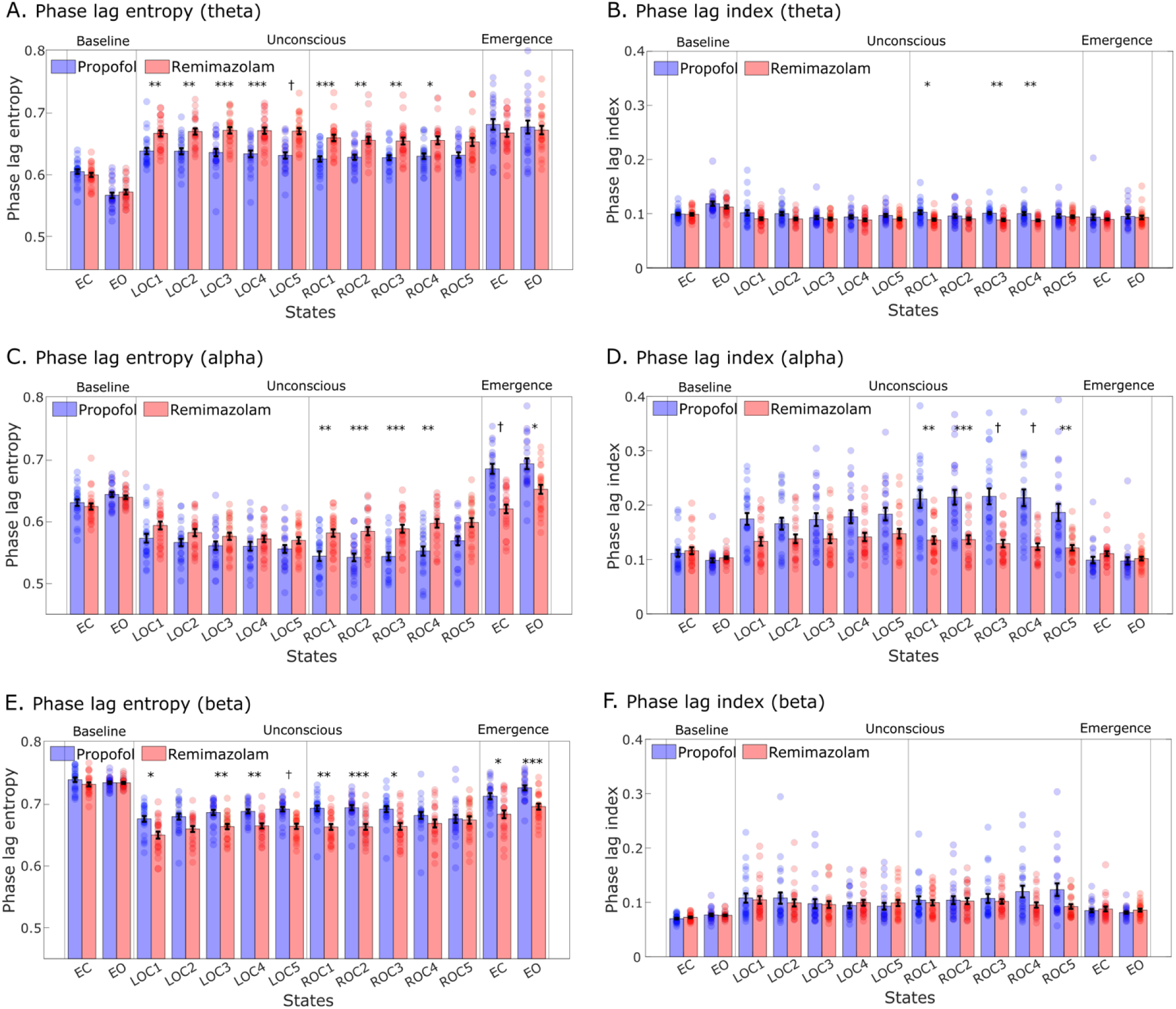
Phase lag entropy (PLE) and phase lag index (PLI) from prefrontal channel pairs. The measures are averaged over all pairs of 4 channels (Fp1, F8, Fp2, and F8) and subjects. Group comparison throughout the baseline to the emergence periods in PLE and PLI at theta (A, B), alpha (C, D), and beta bands (E, F). The error bars represent the standard error for each state. Blue-filled dots represent the individual values from the propofol-induced group (n=24) and red-filled dots represent the individual values from the remimazolam-induced group (n=24). †*P* < 0.0001, \*\*\**P* < 0.001, \*\**P* < 0.01, \**P* < 0.05 in post-hoc analysis.

Conversely, the PLI did not reveal significant differences between the propofol and remimazolam groups in every frequency band upon emergence. During the unconscious period, PLI at the theta frequency band for propofol anaesthesia showed higher values relative to remimazolam anaesthesia in ROC1 (*P* = 0.044), ROC3 (*P =* 0.008), and ROC4 (*P =* 0.007), and PLI at the alpha frequency band showed relatively higher values during the unconscious period compared to the baseline and emergence periods (Figure 3D). Specifically, the propofol group exhibited higher PLI relative to remimazolam anaesthesia across the before-ROC period (*P* = 0.002, *P* < 0.001, *P* < 0.0001, *P* < 0.0001, *P* = 0.006). No group difference was observed at the beta band (Figure 3F). Additional results of connectivity measures at the delta and gamma bands are provided in Supplementary Figure S1.

### Patient state index and correlation among results

The patient state index (PSI) at different time points exhibited a significant group difference during unconscious and emergence periods. Specifically, the propofol group displayed lower PSI values than those of remimazolam group in ROC2 (*P =* 0.008) and ROC3 (*P =* 0.002). Conversely, the PSI values of propofol group were higher than those of remimazolam group during the emergence EC (*P* < 0.0001) and EO (*P =* 0.006) states (Figure 4A). Compared with the baseline, PSI values significantly decreased at all time points after anaesthesia intubation in both anaesthetic agents, and the changes in PSI were greater in the propofol group throughout the unconscious and emergence periods.

**Figure 4.**
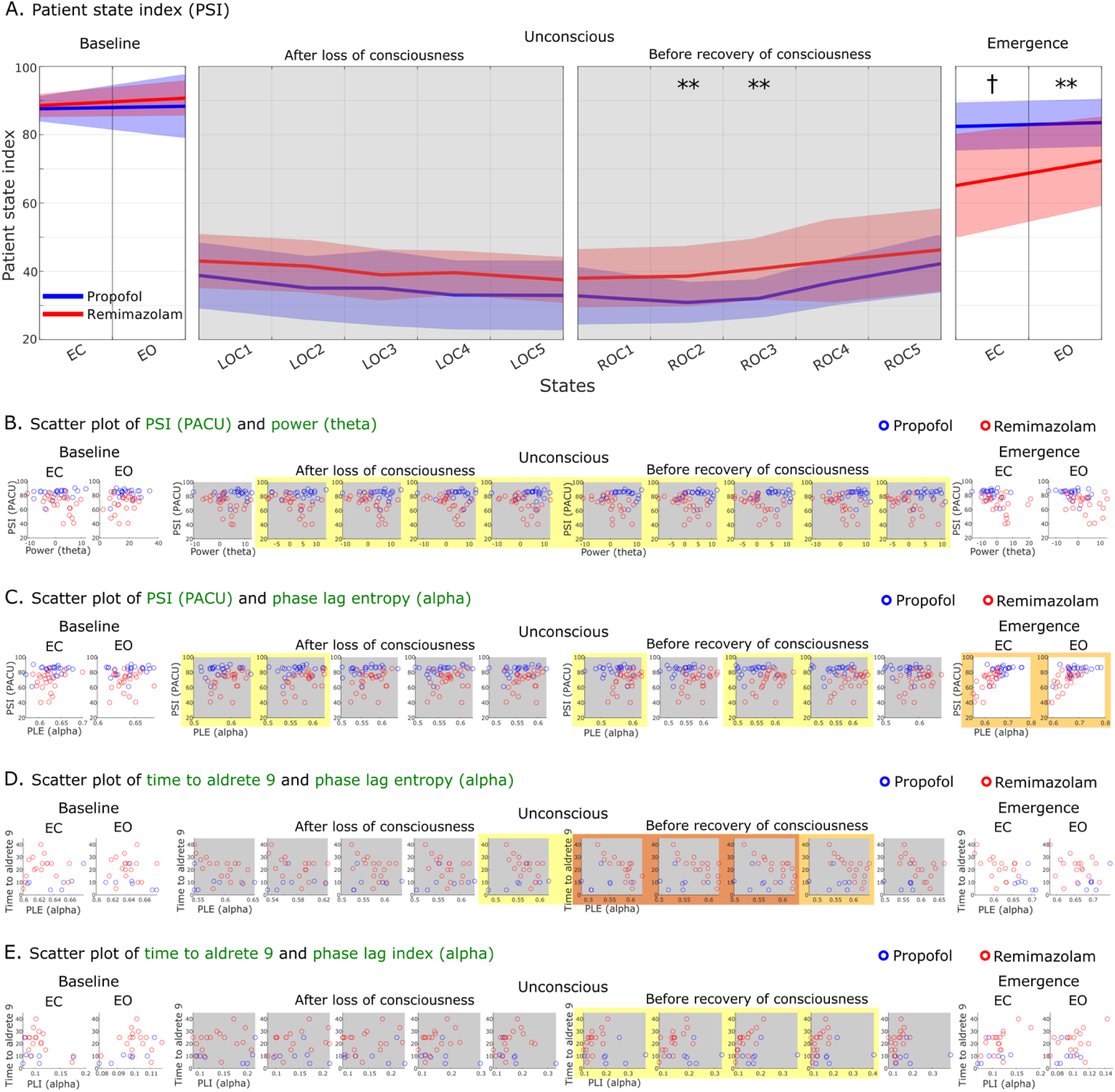
Patient state index (PSI) and comparisons between post-operative recovery indices and EEG measures. (A) Changes in PSI values across baseline, unconscious, and emergence periods in the propofol and remimazolam groups. The ‘after loss of consciousness’ and ‘before recovery of consciousness’ periods are filled with transparent grey. Shaded regions depict standard deviations. †*P* < 0.0001, \*\**P* < 0.01 in post-hoc analysis.

In Pearson’s correlation results, only the results with significant correlation coefficients after Benjamini-Hochberg (FDR) correction (*P* < 0.05) were included in Figure 4B-4E and Table 2, with all significance exclusively observed in remimazolam-induced anaesthesia group. The correlation between recovery indices at PACU (measured using the SedLine system) and EEG measures (functional connectivity and power) throughout the anaesthetic stages revealed both positive and negative correlations. PSI at the PACU showed a negative correlation with theta power (4-8 Hz) during the unconscious period, especially in the LOC2- ROC5 states (Figure 4B and Table 2). Conversely, PSI at the PACU demonstrated a positive correlation with alpha band PLE values during the unconscious and emergence periods, particularly at the LOC1, LOC2, ROC1, ROC3, ROC4, EC, and EO states (Figure 4C and Table 2). However, PLE in theta, beta, PLI in theta, alpha, and beta did not show this correlation.

**Table 2.**
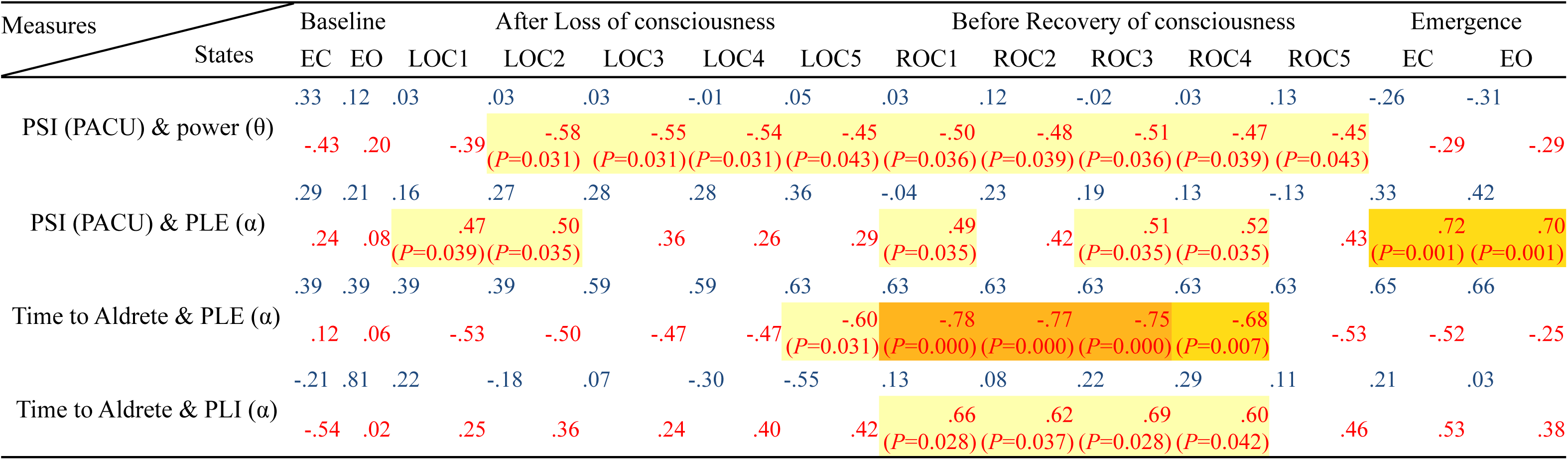
Major Pearson’s correlation among results (FDR-corrected). Correlation coefficients include PSI at PACU and theta power, PSI at PACU and PLE alpha, time to Aldrete 9 and PLE alpha, and time to Aldrete 9 and PLI alpha. The blue and red coefficients represent the correlation values in the propofol and remimazolam groups, respectively. The highlights colored with 3-steps correspond to the significance level of the FDR-corrected *P*-value (yellow: *P* < 0.05, orange: *P* < 0.01, dark orange: *P* < 0.0001).

Comparison between time to Aldrete 9 and two functional connectivity measures exhibited opposite results. A strong negative correlation with PLE alpha was observed in LOC5-ROC4 states during the unconscious period (Figure 4D and Table 2). In contrast, a positive correlation with PLI alpha was observed in the ROC1-ROC4 states during the before-ROC period (Figure 4E and Table 2). Pearson’s correlation results show consistency between the functional connectivity findings in the alpha frequency band and sedation of remimazolam anaesthesia at the PACU (Table 1). Various approaches of PSI and time to Aldrete 9 were observed in correlation analysis (all comparisons, Supplementary Tables S1, S2) and are depicted in scatter plots (Supplementary Figures S2, S3). Analgesia Nociception Index (ANI) scores for each anaesthetic agent and correlations between other result values are provided in Supplementary Figure S4 and Table S3. To demonstrate the prediction of delayed emergence, we utilized both the least squares linear regression and random forest regressor models to predict the time to Aldrete 9. The results of prediction models and the detailed explanations are provided in Supplementary Figure S5.

The scatter plots include significant Pearson’s correlations that survived Benjamini-Hochberg (FDR) correction, such as comparison between (B) PSI values at PACU and theta power, (C) PSI values at PACU and phase lag entropy (alpha), (D) time to Aldrete 9 and phase lag entropy (alpha), (E) time to Aldrete 9 and phase lag entropy (alpha). Excluding subjects with a time to Aldrete 9 value of 0, scatter plots of D & E comprise 7 individuals from the propofol group and 17 from the remimazolam group. The edge highlights colored with 3-steps correspond to the significance level of the FDR-corrected *P*-value (yellow: *P* < 0.05, orange: *P* < 0.01, dark orange: *P* < 0.0001).

## Discussion

The present study highlights three important observations. First and foremost, our results from the PSI values and time to Aldrete 9 at PACU revealed differences between the two anaesthetic agents, indicating slower recovery in remimazolam-induced anaesthesia. Secondly, power values in certain frequency bands differed between the two anaesthetics during and after the anaesthesia. Theta power of remimazolam group throughout the unconscious period was significantly lower compared to propofol group, while higher theta and alpha power was observed under emergence period. Third, differences in functional connectivities between groups were observed, with remimazolam-anaesthesia exhibiting higher PLE in the theta and alpha band during unconsciousness, whereas lower PLE in the alpha and beta bands during emergence, and higher PLI in the alpha band during unconsciousness. Additionally, PLE and PLI at the alpha band under remimazolam-induced unconsciousness were highly correlated with time to Aldrete 9 at PACU. This not only corroborates the notion of previous studies (i.e. Lee and colleagues^11^) suggesting a link between anaesthetically induced unconsciousness and reduction of PLE, but also indicates that PLE and PLI at the alpha band may provide insight into the speed of emergence in patients under remimazolam-induced unconsciousness.

In summary, dynamic changes in the power and functional connectivity in theta and alpha bands during the transition from unconsciousness to wakefulness significantly elucidate the distinct prefrontal EEG features between the two anaesthetics. These findings offer valuable insights into predicting how patients’ levels of consciousness vary upon emergence from anaesthesia with the two different agents.

Our correlation results, exclusively observed with remimazolam-induced anaesthesia, suggest that the EEG patterns during remimazolam-induced unconsciousness may predict the emergence speed of remimazolam anaesthesia. Specifically, the negative correlation between PSI scores at PACU and theta power during unconsciousness accounts for the relationship between the level of consciousness and the degree of theta power. High theta power (< 10 dB) during unconsciousness is associated with low consciousness levels at PACU, while low theta power (< -10 dB) is related to high consciousness levels. Conversely, the positive correlation between PSI at PACU and PLE at the alpha band under unconscious and emergence suggests that reduced PLE values during emergence correspond to delayed recovery of consciousness at PACU.

More importantly, time to Aldrete 9 exhibited strong correlations with PLE and PLI at the alpha band during before-ROC period, though not shown upon emergence. The negative correlation between time to Aldrete 9 and PLE alpha showed that slower achievement of Aldrete score 9 is associated with reduced diversity of phase patterns during anaesthesia. The positive correlation between time to Aldrete 9 and PLI alpha before ROC showed that slower achievement of Aldrete score 9 is associated with increased phase lead/lag relationship during anaesthesia. Moreover, the results of least square linear regression and random forest regression further strengthen the correlation with time to Aldrete 9 and support that alpha band PLE and PLI during the unconscious state may predict emergence speed under remimazolam anaesthesia, potentially due to enhanced frontal alpha oscillations mediated by potentiated GABA receptors^15, 18^. The prediction of emergence speed in remimazolam-induced anaesthesia can as well impact the efficient utilization of flumazenil.

The recovery from remimazolam anaesthesia displayed delayed awakening, as indicated by PSI and time to Aldrete 9 in the PACU. Prefrontal EEG profiles during emergence showed distinct characteristics for remimazolam versus propofol, possibly contributing to the delayed awakening observed with remimazolam. Previous studies have suggested that this delayed emergence may be due to changes in drug concentrations or drug interactions, distinct neurobiological mechanisms between the anaesthetics, or differences in depth of anaesthesia monitoring systems. Literature indicates that the combination of remifentanil and propofol induces faster emergence from propofol anaesthesia compared to remimazolam anaesthesia.^20^ On the other hand, remifentanil and remimazolam interaction may influence delayed emergence.^21, 22^

Furthermore, previous studies suggest that lower remimazolam dosages facilitate shorter recovery^8, 23^, while higher dosages may prolong it.^2, 4^ A target range of anaesthesia depth also affects emergence time; a BIS value of 50-59 reduces propofol consumption and shortens emergence compared to a BIS value of 40-49.^24^ In most of the previous studies comparing anaesthetic depth between propofol and remimazolam anaesthesia using Bispectral Index (BIS) or SedLine monitoring, consistent with our findings, higher remimazolam values were observed compared to propofol anaesthesia during the maintenance of anaesthesia regardless of the dosage level^2, 4, 6, 23^, with an exception noted in one study^25^. While many studies have compared anaesthetic depth between propofol and remimazolam, few have focused on EEG patterns of dynamic changes in consciousness during emergence^2, 4, 6, 8, 23, 25^ (Table S5). Monitoring with PSI appears more sensitive to rapid pharmacokinetic changes in remimazolam compared to BIS, likely due to algorithmic differences.^26–28^ Unlike BIS, SedLine monitoring separates EEG and EMG signals, reducing muscle activity interference.^29^ Factors like remifentanil and remimazolam interaction, dosage levels, anaesthesia depth monitoring systems, and flumazenil administration impact remimazolam-induced emergence speed.

Our research holds the advantage of being the first to explore short-term delayed emergence in remimazolam-induced anaesthesia through dynamic changes in the level of consciousness using EEG power and functional connectivity approaches. Our results highlight the necessity for management strategies addressing short-term delayed emergence at the PACU. However, several factors warrant further investigation. First, to capture a more comprehensive picture of changes in consciousness levels, extending the duration of PSI measurement at the PACU, as demonstrated by Doi and colleagues2, would allow us to identify the point at which the consciousness levels under the two anaesthetic agents converge. Second, exploring drug-drug interactions with opioids other than remifentanil could provide further insights into how remimazolam and propofol differently interact with remifentanil and other opioids. Through these additional investigations, we may gain insights into more robust picture of delayed arousal and changes of consciousness level, and the impact of remimazolam-remifentanil interaction on emergence speed.

Due to the faster recovery of consciousness in propofol-induced anaesthesia compared to remimazolam-induced anaesthesia, a significant number of subjects had already attained Aldrete 9 when assessing consciousness levels at PACU. Consequently, determining whether there exists a correlation between EEG correlates and emergence speed in propofol was challenging. However, by measuring consciousness levels at narrower intervals starting from the termination of anaesthetic in future experiments, it might be feasible to validate whether these findings also extend to propofol-induced anaesthesia.

## Supporting information

Supplementary Materials Combined

## Data Availability

All data produced in the present study is not open and is not available to the public.

## Author’s contribution

KBN, MJY: Study design

KBN: Ethics approval and registration

KBN, PSJ, KJM, KEJ: Patient recruitment and data collection

LYJ, KHK, PSJ, PYJ, KJW, MJY: Data analysis

LYJ, KHK, LUC, MJY, KBN: drafting

## Acknowledgements

We thank the Institute of Basic Science and National Research Foundation of Korea for funding this work. We thank Heonsoo Lee for insightful advices on the analysis. We thank all the patients for kindly participating in this work.

