## Supplementary Materials Combined for "EEG Correlates of Delayed Emergence after Remimazolam-Induced Anaesthesia Compared to Propofol"

**Supplementary Material**

*Study Procedure*

A computer-generated randomization table (available at https://www.randomizer.org/form.htm) was used to assign patients to the remimazolam or propofol group at a 1:1 ratio. Among the 60 patients enrolled in this study, individuals with low PSI scores at baseline, insufficient EEG data, or a lack of marker flag were excluded. Consequently, we analyzed the data from the remaining 48 patients (n = 24 per group) (Figure 1A). Patients, operators, and two research personnel were unaware of the group identity, but the attending anesthesiologist couldn't be blinded about the group identity due to the distinct properties and color of the two anesthetics.
Before entering the operating rooms, we placed the SEDLine sensor on the patient's forehead. Preoperative baseline EEG were recorded at controlled eye-open (focused wakefulness) and eye-closed (relaxed wakefulness) conditions, for each 4-minute period.

Upon entering the operating rooms, the patients were monitored with pulse oximetry, non-invasive arterial blood pressure, electrocardiography, and anesthetic depth (SedLine®, Masimo Corp., Irvine, CA, USA). The patients received 0.1 mg glycopyrrolate before infusing remifentanil and remimazolam or propofol using a commercial syringe pump (Agillia, SB Medica SRL, Italy).

In the propofol group, anesthesia was induced using propofol [target-controlled infusion (TCI), Marsh model] and remifentanil at effect-site concentrations of 4 mcg $ml^{-1}$ and 3 ng $ml^{-1}$ respectively. The concentration of propofol is adjusted to maintain an appropriate depth of anesthesia based on the EEG (PSI target 40) until the end of the surgery. In the remimazolam group, anesthesia was induced using remifentanil at an effect-site concentration of 3 ng $ml^{-1}$ (TCI, Minto model) and remimazolam at a flow rate of 6 mg $kg^{-1}$ $h^{-1}$, as per the manufacturer's recommendations. The concentration of remimazolam is adjusted to maintain an appropriate depth of anesthesia based on the EEG (PSI target 40) until the end of the surgery.

In both groups, neuromuscular blockade was induced using intravenous rocuronium (1 mg $kg^{-1}$) after loss of consciousness. At 3 minutes after rocuronium administration, endotracheal intubation was attempted using a video laryngoscope and an endotracheal tube in both groups. Patients in both groups received 0.3 mg ramosetron for postoperative nausea and vomiting and 1mcg $kg^{-1}$ fentanyl on the skin closure for pain control. All the patients received sugammadex for neuromuscular block reversal.

EEG at the post-anesthesia care unit was also recorded at controlled eye-open and eye-closed conditions, at the time when the patient could respond to commands, for each 4-minute period. Patients who have successful emergence from the ventilator with spontaneous eye-opening were moved to the Post-Anaesthesia Care Unit (PACU). Sedation and agitation were promptly evaluated using the Richmond Agitation-Sedation Scale (RASS) immediately upon admission to PACU (Table S4). The Aldrete score, a measure of fitness for discharge from the PACU, was also measured immediately upon admission to the PACU and assessed every 2 minutes. EEG at the post-anaesthesia care unit was also recorded at controlled eye-open and eye-closed conditions, at the time when the patient could respond to commands, for each 4-minute period. Pain was assessed using the Numeric Rating Scale (between 0 and 10,0 = no pain,10 = the most severe pain). If NRS > 5, rescue analgesic (fentanyl) was administered intravenously.

The primary outcome was frontal spectral power in EEG (Electroencephalography), which refers to the measurement of electrical activity in the frontal lobes of the brain across different frequency bands. Frontal spectral power specifically focuses on the electrical activity in the frontal region of the brain within these frequency bands. Different frequency bands are associated with different states of brain activity. For example, alpha waves are often associated with relaxation or idling of the brain, while beta waves are linked to more active cognitive processing. The secondary outcome was RASS score at PACU and the time to reach an Aldrete score of at least 9 in PACU. The Aldrete score assesses post-anesthesia recovery based on various criteria, including mobility, respiration, oxygenation, cardiovascular stability, and consciousness. The Aldrete score of 9 is commonly considered satisfactory for discharge to general ward.

### *I. Power Spectrogram and Line Power Analyses*

Power spectrogram was computed to compare the frequency power during unconsciousness and after emergence from anesthesia between the two groups. We used "pspectrum.m" function in the MATLAB Toolbox which analyze signals in the frequency and time-frequency domains. To compare the power spectrogram of the two anesthetic drugs at a freqeuncy over time, 1,024 frequency bins were obtained in the frequency domain by dividing the range of 1-50Hz into intervals of 0.0479 Hz. The time window size was set as 3 seconds using a Hann window (leakage factor for a kaiser window = 0.85) with 50% overlap, resulting in a size of 1,680 time bins. By considering both time and frequency, a total of 1,720,320 (1,024 x 1,680) group comparisons are possible. Each power value was converted to decibel (dB) units [$20 \times \log_{10}$ (amplitude)]. The power dB ratio between the two anesthetic agents was then obtained by dividing remimazolam dB by propofol dB within a frequency range of 1-50 Hz across all times (min).

$$\text{Calculationofpowerratio}\,(1-50\text{Hz}) = \frac{\text{Remimazolampower}\,(\text{dB})}{\text{Propofolpower}\,(\text{dB})}$$

Propofol dB greater than remimazolam dB was indicated as blue, whereas remimazolam dB greater than propofol dB was indicated as red. To statistically represent the power difference between the two different anesthetic agents over time in a specific frequency range, spectral power was integrated to obtain each power dB corresponding to each of the five frequency bands.

### *II. Functional Connectivity Measure Analysis*

Two phase measures were applied to the EEG data analysis (sampling rate 178 Hz) by upsampling the data to 200 Hz. Specifically, we applied phase lag entropy (PLE) and phase lag index (PLI) at the frequency bands identical to those in the power analyses. We calculated PLE and PLI within uniformly segmented 10-sec epochs which was then averaged when comparing between the two groups. With these applied measures, the diversity of the entropy pattern (PLE), the degree of phase lead/lag synchronization and the phase locking (PLI) between the prefrontal EEG signals are calculated. These measure analyses assess the group difference in the phase relationship between the two prefrontal EEG signals following loss of consciousness induced by different anesthetic agents. This was performed by averaging four channels. A crucial difference between phase synchronisation measure (PLI) and PLE is that the temporal pattern of the phase relationships between the two signals can be extracted from PLE, while PLI can only calculate the mean phase locking.[1]

**Phase lag entropy**

Phase lag entropy (PLE) is a phase synchronization measure used in Lee's study[1], which was suggested as a hypnotic depth indicator. PLE measures the temporal pattern diversity of phase lag difference between the two signals, where this information allows us to detect how neural communication changes across time. The steps of PLE calculation follow Lee's approach. First step is to extract instantaneous phase signals from the two signals, which can be extracted through Hilbert transform using the signal processing toolbox in MATLAB (version R2022b, Mathworks Inc., Co., Ltd., USA). Second, symbolize the phase difference of the signals in two conditions, $s_t = 0$ if phase difference is less than 0, $\Delta\phi_t < 0$ (first signal is the phase lag) and $s_t = 1$ if phase difference is greater than 0, $\Delta\phi_t > 0$ (the first signal is the phase lead). Then, the temporal pattern of phase relationship, $S_t$, is modulated by parameters *m* and τ, where *m* depicts the size of the pattern and τ depicts the time lag.

$$S_t = \{s_t, s_{t+\tau}, \cdots s_{t+(m-1)\tau}\}, t = 1,2,\ldots,N-(m-1)\tau,$$

In our analysis, we used *m* = 3 (generating 8 patterns: "000", "001", "010", "100", "011", "101", "110", and "111") and τ= 3. Lastly, standard Shannon entropy formula is applied to the phase patterns which allows estimating the number of pattern occurrences in the given epoch.

$$\mathrm{PLE} = \frac{-1}{\log(2^m)} \sum_j p_j \log(p_j),$$

From the PLE equation, $p_j$ indicates the probabiility that the $j^{th}$ pattern will occur at a given input signal. The numerator represents the entropy of probability of obtaining different phase patterns in the signal, and the denominator represents the number of achievable patterns. The denominator scale of the PLE values is ranged between 0 and 1, where values close to 0 (PLE ≈0) indicates the simplicity of patterns, thus lead/lag relationship is predictable, whereas values close to 1 (PLE ≈1) indicates the diversity of patterns, signifying unpredictable lead/lag relationship.[1]

**Phase lag index**

Phase lag index is a widely used method in EEG and magnetoencephalogram data as it measures the averaged phase locking and the degree of phase lead/lag synchronization between the two signals. A basic concept of PLI suggests that when there are consistent nonzero phase lags between two signals, it represents genuine interaction, not due to a volume conduction. As in PLE, instantaneous phase signals from the two signals are calculated via Hilbert transform.

$$\mathrm{PLI} = \frac{1}{\mathrm{N}}\left|\sum_{t=1}^{\mathrm{N}} \mathrm{sign}(\Delta\phi_t)\right|,$$

From the equation above, the sign function gives 1 if phase difference is greater than 0, $\Delta\phi_t > 0$; whereas the sign function gives -1 if phase difference is less than 0, $\Delta\phi_t < 0$. PLI values close to 0 (PLI $\approx$0) indicates an absence of (zero) phase lag synchronization or severe volume conduction; whereas PLI values close to 1 (PLI $\approx$1) indicates the presence of phase lagged synchronization or complete-locking with nonzero phase lag.[2]

**Regression Analyses**

We conducted regression analysis based on features obtained during surgery to predict the time to Aldrete 9. We utilized the features that exhibited high correlation in the correlation analysis for this purpose. The features were derived from the average value of Alpha PLE before ROC 1, 2, and 3. We compared two regression models: least squares linear regression and random forest regression [3]. Considering the limited dataset size, leave-one-out cross-validation (LOOCV) was employed to assess the models' performance. The evaluation metric used for assessing model performance was the mean absolute error (MAE). The analysis was carried out using the Python programming language along with the Scikit-learn library[4].

A. Phase lag entropy (delta)

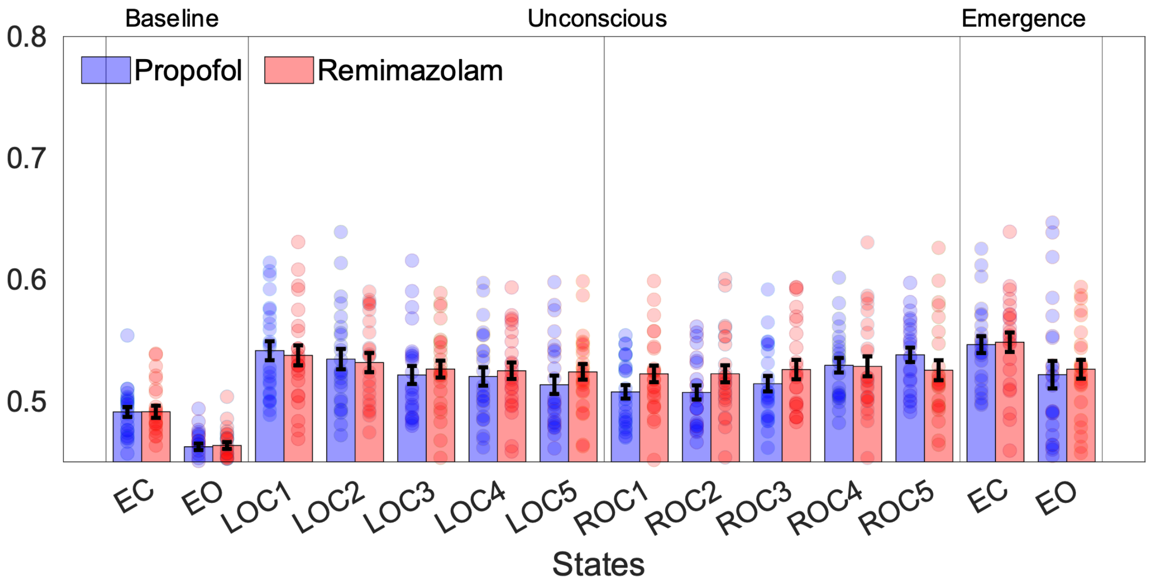


B. Phase lag index (delta)

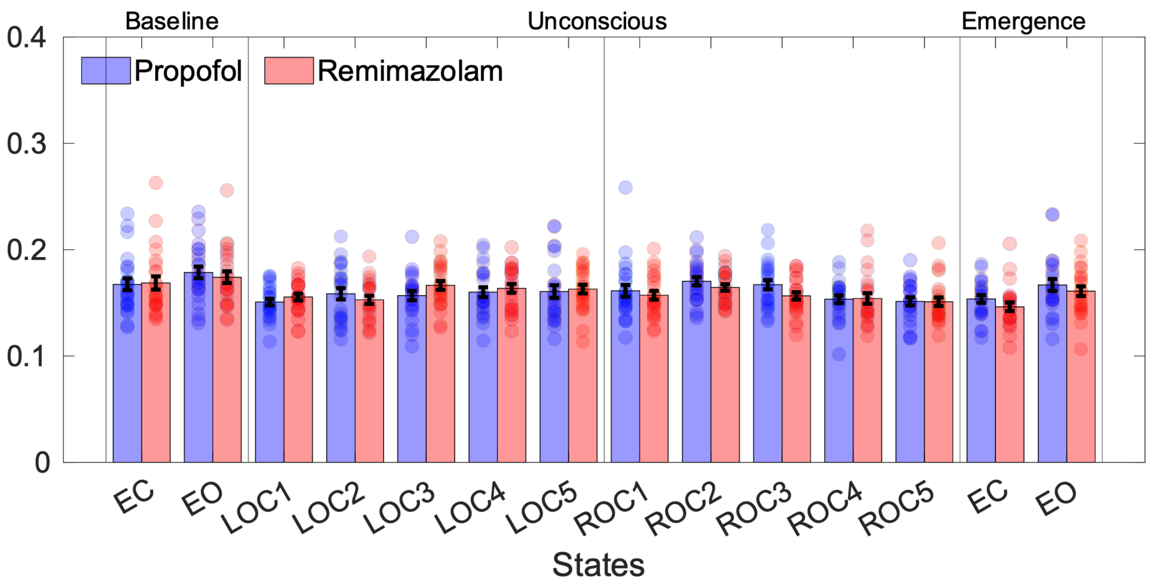

C. Phase lag entropy (gamma)

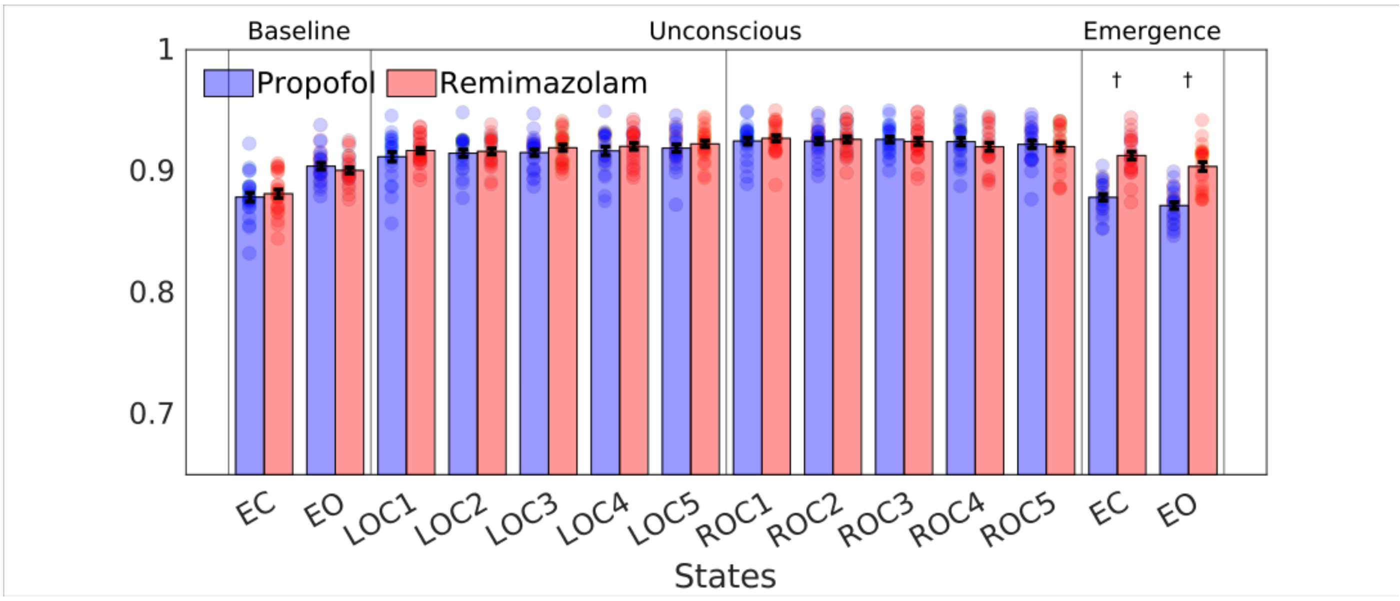


D. Phase lag index (gamma)

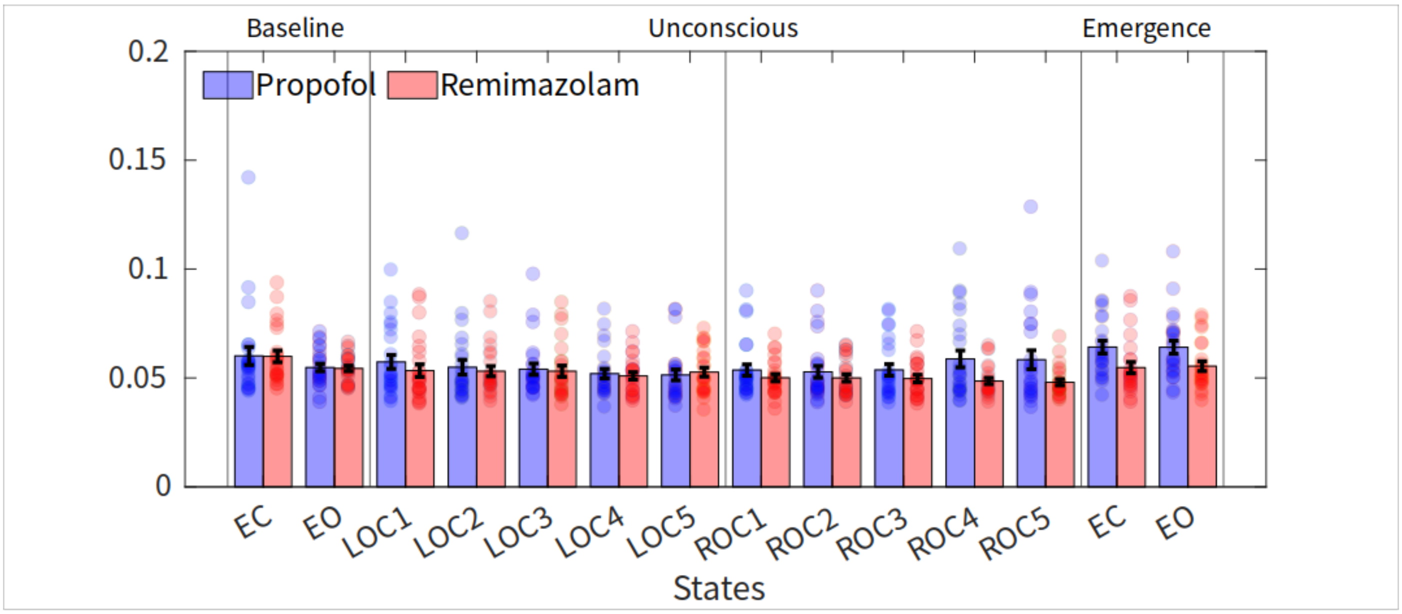


**Figure S1.** Functional connectivity measures. Phase lag entropy (PLE) and phase lag index (PLI) from prefrontal-frontal channel pairs at delta and gamma bands. PLE and PLI at the beta frequency band showed no group difference during any anesthesia period (A, B). On the other hand, PLE at the gamma frequency band showed a significant group difference during emergence from the anesthesia (C). However, PLI at the gamma band showed no change in phase synchronization from the baseline throughout the anesthesia process, and no difference between groups (D). †$P < 0.0001$ in post-hoc analysis.

A. PSI and functional connectivity

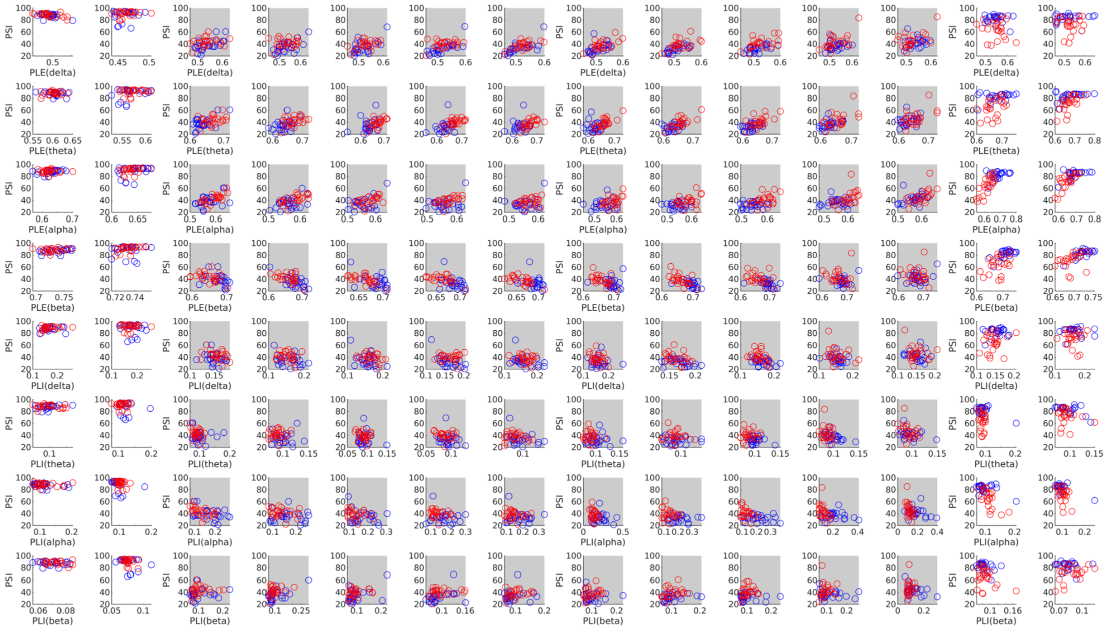


B. PSI and powers

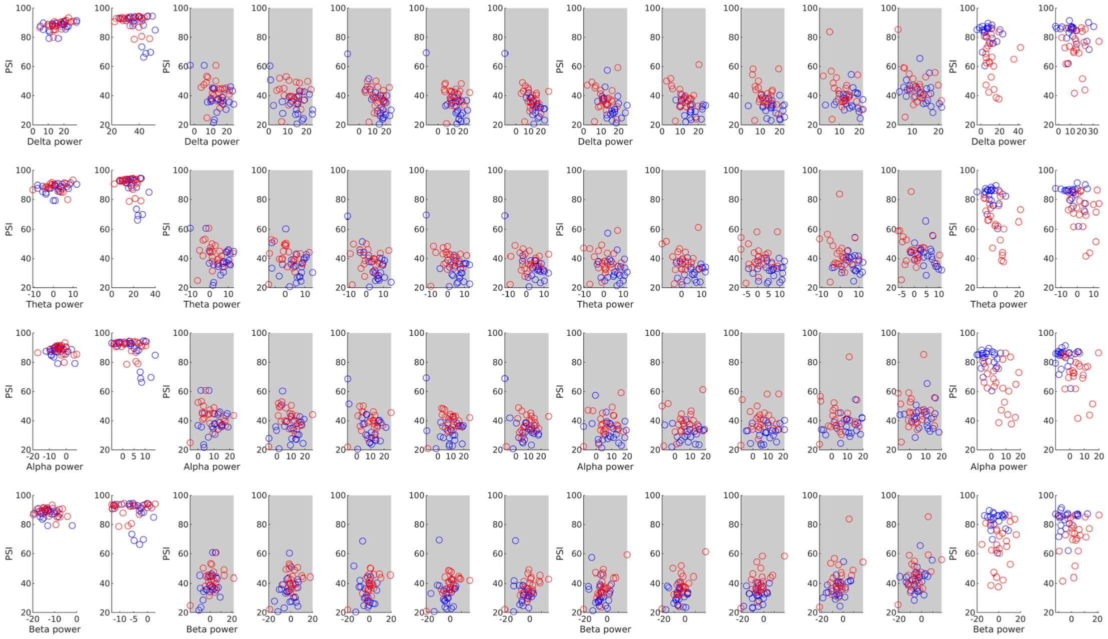

C. PSI (PACU) and functional connectivity

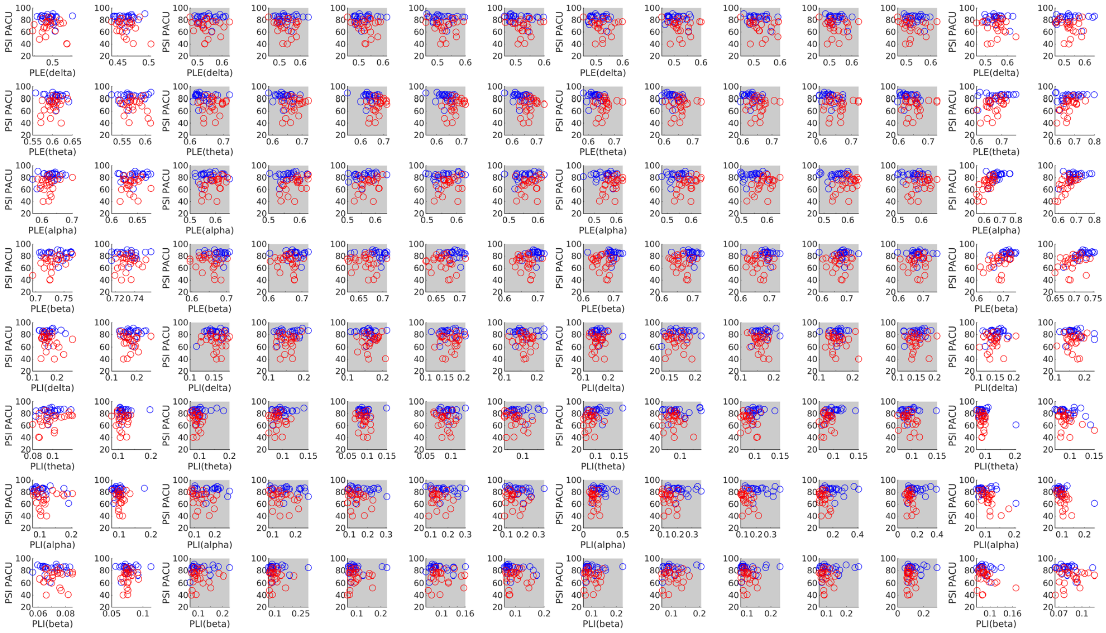


D. PSI (PACU) and analysis results

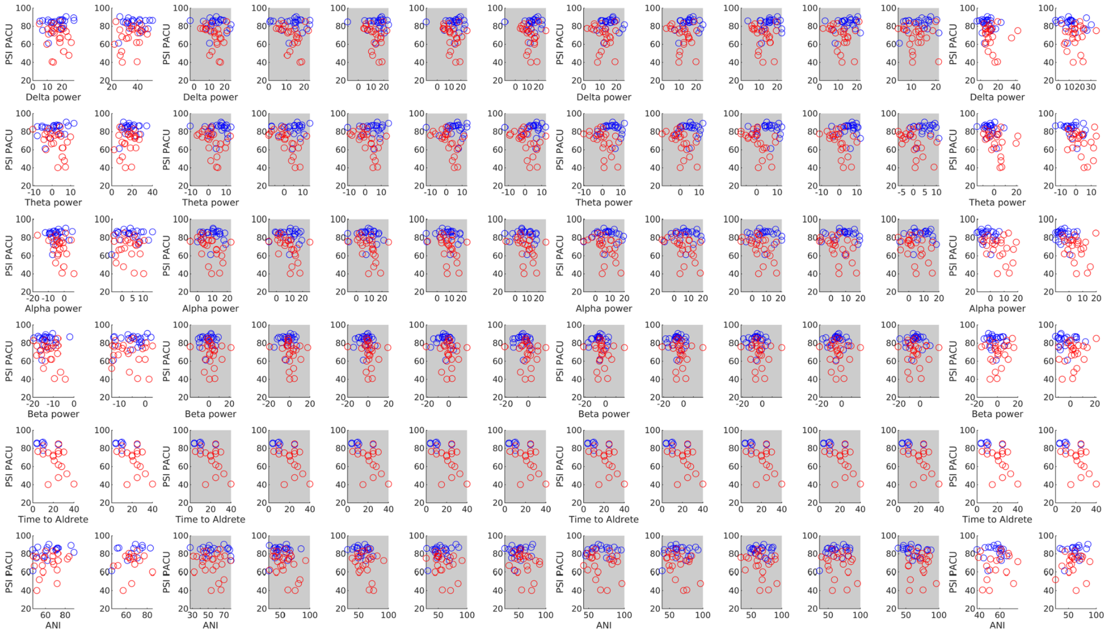


**Figure S2.** Scatter plots of comparisons between PSI values and measure values throughout the anesthesia process.

Comparisons include (A) PSI scores and functional connectivity (PLE and PLI at four frequency bands), (B) PSI scores and powers at four frequency bands (delta, theta, alpha, and beta), (C) PSI scores at PACU and functional connectivity (PLE and PLI at four frequency bands), and (D) PSI scores at PACU and analysis results including four powers, time to Aldrete 9, and ANI scores. The blue and red scores represent the correlation values in propofol and remimazolam groups, respectively. *ANI = analgesia nociception index

A. Time to Aldrete 9 and functional connectivity

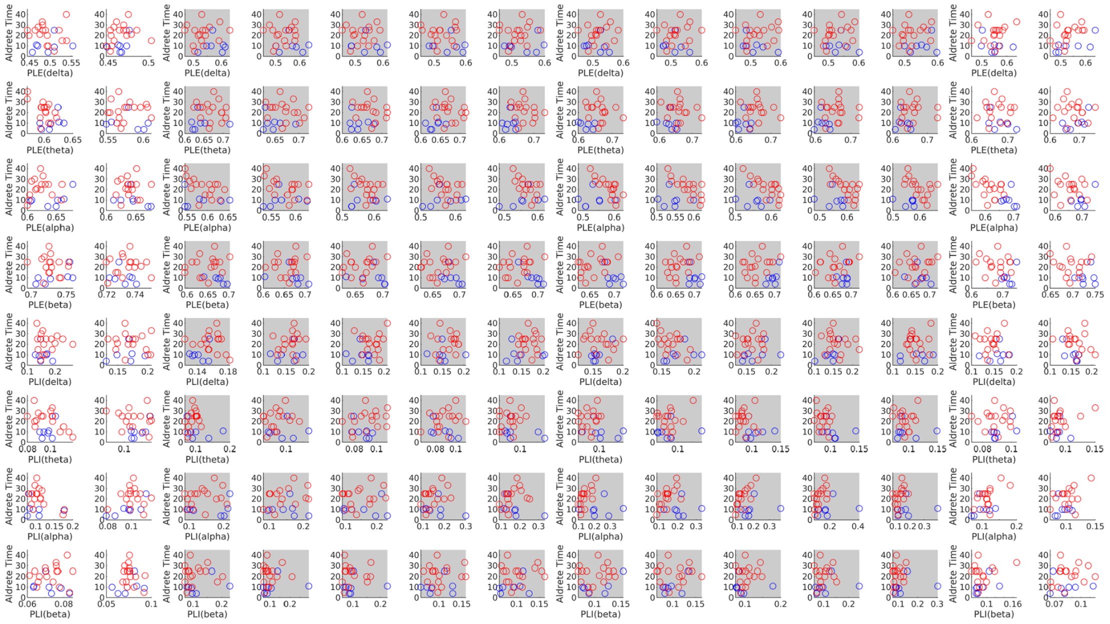

B. Time to Aldrete 9 and analysis results

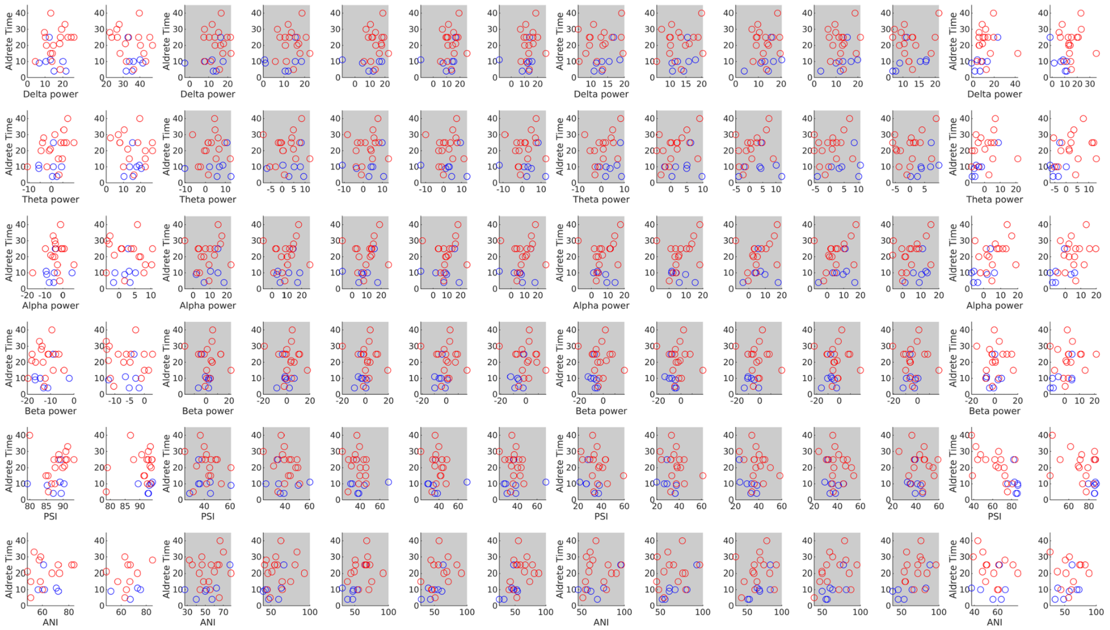

**Figure S3.** Scatter plots of time to Aldrete 9 and analysis results across all anesthesic periods. Scatter plots in (A) represent a comparison between time to Aldrete 9 (min) and PLE or PLI at four frequency bands. Patients taken zero minute to reach Aldrete 9 were excluded (leaving 7 propofol, 17 remimazolam). Scatter plots in (B) represent a comparison between time to Aldrete 9 (min) and analysis results including four powers (delta, theta, alpha, and beta), PSI, and ANI scores across all anesthetic periods.

A. Analgesia Nociception Index (ANI)

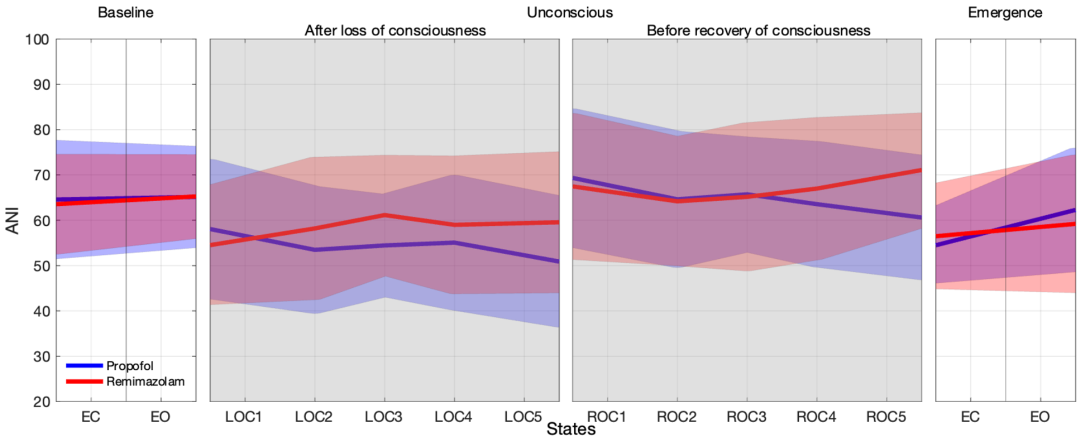


B. ANI and functional connectivity

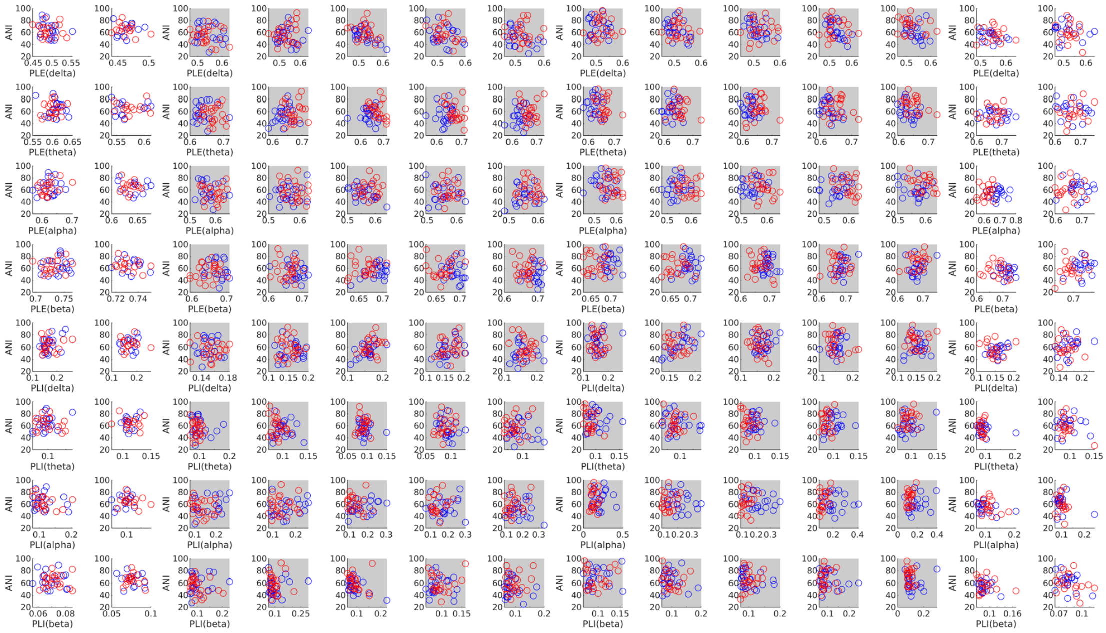

### C. ANI and analysis results

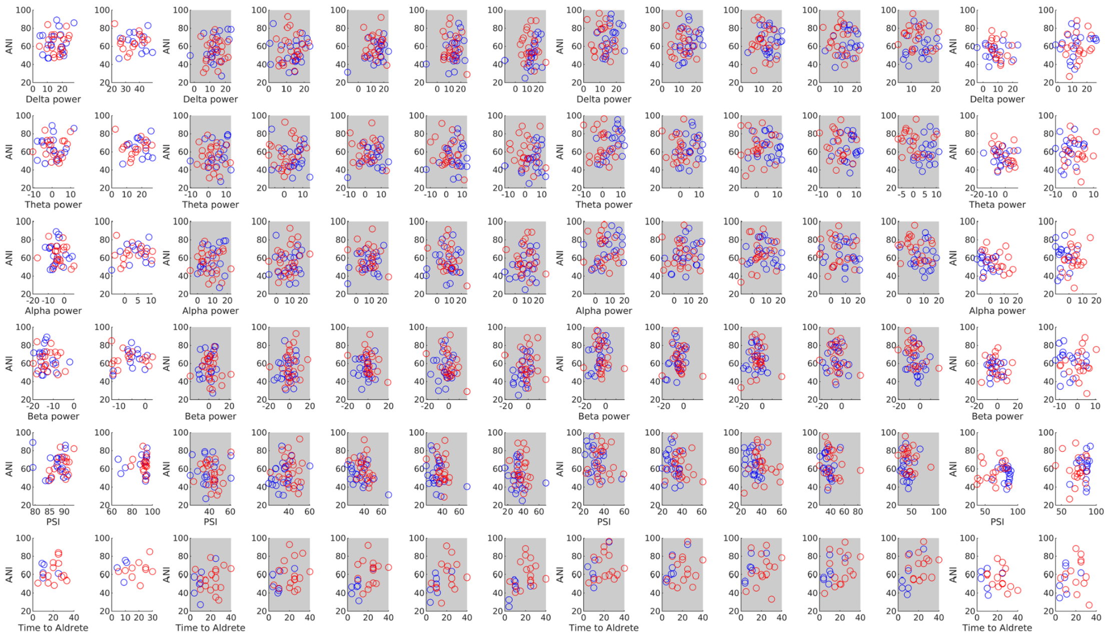


### D. ANI (PACU) and functional connectivity

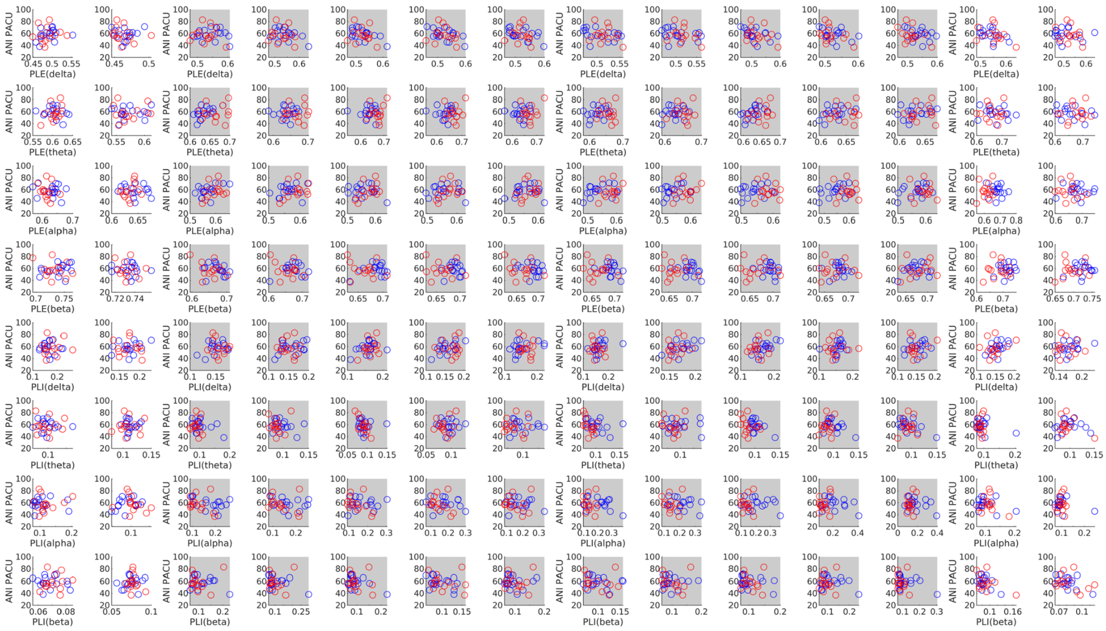

E. ANI (PACU) and analysis results

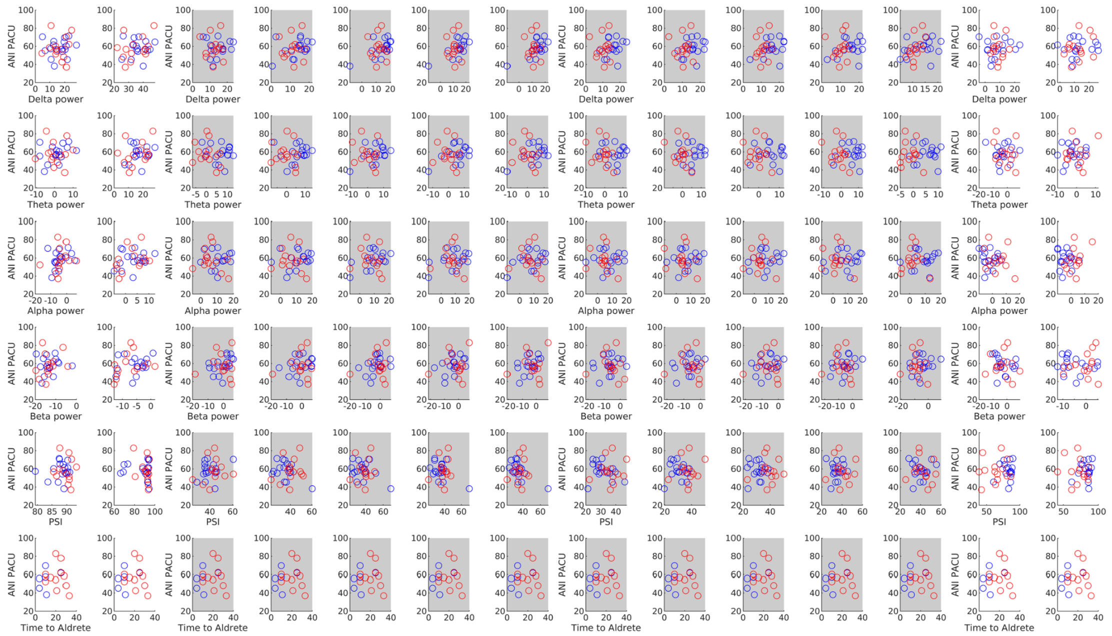


**Figure S4.** ANI values and comparison between ANI values and measure values.

(A) Changes in analgesia nociception index (ANI) values for each group during baseline (EC, EO), after loss of consciousness, before recovery of consciousness, and emergence (EC, EO) periods. The ANI values of remimazolam and propofol showed insignificant differences throughout the anesthetic process. Scatter plots in (B) represent a comparison between ANI and functional connectivity (PLE and PLI) at four frequency bands (delta, theta, alpha, and beta) across all anesthesia periods. Scatter plots in (C) represent a comparison between ANI and analysis results including powers at four frequency bands (delta, theta, alpha, and beta), PSI, and time to Aldrete 9 values across all anesthetic periods. Scatter plots in (D) depict a comparison between ANI values at PACU and functional connectivity at four frequency bands across all anesthesia periods. Scatter plots in (E) depict a comparison between ANI values at PACU and analysis results including powers at four bands, PSI, and time to Aldrete 9 values across all anesthetic periods.

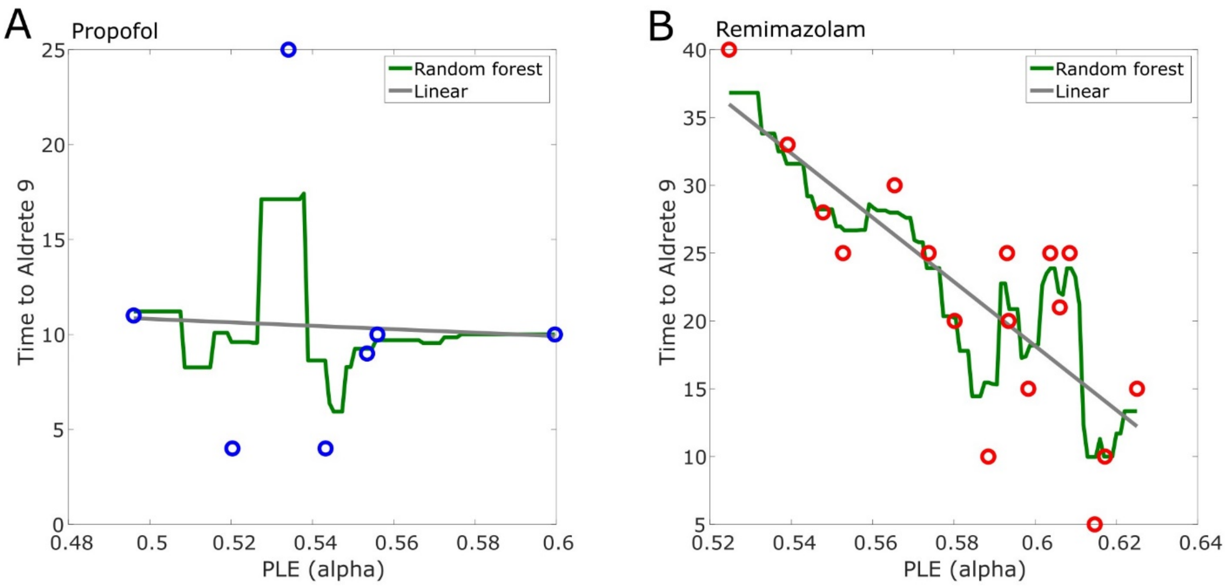


**Figure S5.** Prediction of delayed emergence.

Regression analysis was conducted to forecast delayed emergence. We utilized both the least squares linear regression (A) and random forest regressor models (B) to predict the time to Aldrete 9. The performances of these models were assessed using leave-one-out cross-validation (LOOCV), and the mean absolute errors were calculated (details included in the Supplementary method section, Regression analyses). When computed with the Random Forest Regressor, the MAE was 4.89, whereas with Linear Regression, the MAE was 5.03. Regression results based on the average alpha band PLE values of ROC 1, 2, and 3 predicted a time to Aldrete 9 that differed by approximately 5 minutes from the actual value (Table S6).

A. Correlation coefficients (N=48)

| Measures \ States | Baseline | | After Loss of consciousness | | | | | Before Recovery of consciousness | | | | | Emergence | |
|---|---|---|---|---|---|---|---|---|---|---|---|---|---|---|
| | EC | EO | LOC1 | LOC2 | LOC3 | LOC4 | LOC5 | ROC1 | ROC2 | ROC3 | ROC4 | ROC5 | EC | EO |
| PSI & PLE (δ) | -.47 | .19 | .58 | .62 | .57 | .73 | .70 | .48 | .72 | .65 | .21 | .16 | -.12 | .08 |
| | -.61 | .20 | .23 | .11 | .43 | .29 | .62 | .63 | .54 | .51 | .58 | .51 | -.17 | -.04 |
| PSI & PLE (θ) | -.23 | .32 | .57 | .43 | .36 | .35 | .23 | .05 | .41 | .33 | .45 | .30 | .37 | .42 |
| | .24 | .14 | .33 | .42 | .62 | .59 | .62 | .75 | .73 | .59 | .54 | .55 | .39 | .62 |
| PSI & PLE (α) | .46 | .36 | .43 | .41 | .43 | .22 | .24 | .15 | .13 | .19 | .07 | .29 | .33 | .48 |
| | -.05 | .28 | .72 | .62 | .48 | .54 | .20 | .43 | .36 | .44 | .50 | .62 | .79 | .73 |
| PSI & PLE (β) | .44 | .09 | -.32 | -.64 | -.42 | -.38 | -.34 | -.04 | -.50 | -.37 | .09 | .25 | .63 | .68 |
| | .19 | .35 | -.40 | -.47 | -.67 | -.61 | -.67 | -.56 | -.58 | -.38 | -.12 | -.16 | .39 | .38 |
| PSI & PLI (δ) | .05 | -.15 | -.27 | -.34 | -.43 | -.64 | -.36 | -.11 | -.38 | -.39 | -.29 | -.17 | .11 | -.06 |
| | .36 | .07 | -.32 | -.14 | -.28 | .08 | -.16 | -.34 | -.32 | -.39 | -.29 | -.17 | .21 | -.01 |
| PSI & PLI (θ) | .41 | -.18 | -.08 | .03 | -.08 | -.20 | -.18 | .00 | -.21 | -.17 | -.44 | -.34 | -.61 | -.60 |
| | -.09 | .01 | -.34 | .04 | -.17 | -.25 | -.04 | -.07 | -.02 | -.01 | -.15 | -.36 | -.14 | -.17 |
| PSI & PLI (α) | -.32 | -.22 | -.35 | -.03 | -.26 | -.05 | -.07 | -.12 | -.15 | -.19 | -.10 | -.26 | -.44 | -.68 |
| | -.04 | -.24 | -.46 | -.24 | -.18 | -.17 | -.09 | -.23 | -.13 | -.36 | -.43 | -.48 | -.66 | -.46 |
| PSI & PLI (β) | -.14 | -.25 | .08 | .60 | .44 | .39 | .18 | -.05 | .31 | .01 | -.14 | -.32 | -.35 | -.15 |
| | -.26 | -.29 | .16 | .30 | .28 | .20 | .30 | .27 | .33 | .23 | .11 | .14 | -.36 | .31 |
| PSI & power (δ) | .39 | -.46 | -.38 | -.46 | -.80 | -.75 | -.81 | -.38 | -.33 | -.32 | -.07 | -.40 | -.03 | -.16 |
| | .54 | -.30 | -.20 | -.01 | -.12 | -.06 | -.21 | -.17 | -.22 | -.24 | -.41 | -.34 | -.05 | -.01 |
| PSI & power (θ) | .28 | -.49 | -.30 | -.19 | -.71 | -.57 | -.68 | -.20 | -.04 | .01 | .01 | -.36 | -.23 | -.29 |
| | .04 | -.28 | -.13 | -.15 | -.04 | -.01 | .07 | .02 | -.09 | -.03 | -.17 | -.17 | -.25 | -.25 |
| PSI & power (α) | -.17 | -.54 | .04 | -.05 | -.49 | -.30 | -.48 | -.22 | .11 | .23 | .28 | -.04 | -.13 | -.02 |
| | -.18 | -.16 | -.16 | -.08 | .11 | .21 | .34 | .26 | .16 | .17 | .10 | .06 | -.33 | -.26 |
| PSI & power (β) | -.36 | -.25 | .37 | .18 | -.21 | -.09 | -.36 | -.18 | .14 | .28 | .37 | .27 | -.08 | .17 |
| | -.28 | .14 | .35 | .38 | .61 | .62 | .60 | .65 | .61 | .53 | .47 | .51 | .21 | .26 |

B. Benjamini-Hochberg (FDR) corrected *P*-values (N=48)

| Measures / States | Baseline | | After Loss of consciousness | | | | | Before Recovery of consciousness | | | | | Emergence | |
|---|---|---|---|---|---|---|---|---|---|---|---|---|---|---|
| | EC | EO | LOC1 | LOC2 | LOC3 | LOC4 | LOC5 | ROC1 | ROC2 | ROC3 | ROC4 | ROC5 | EC | EO |
| PSI & PLE (δ) | 0.030 | 0.482 | 0.006 | 0.003 | 0.006 | 0.000 | 0.000 | 0.030 | 0.000 | 0.003 | 0.451 | 0.530 | 0.614 | 0.726 |
| | 0.009 | 0.431 | 0.378 | 0.668 | 0.067 | 0.266 | 0.007 | 0.007 | 0.017 | 0.022 | 0.011 | 0.022 | 0.501 | 0.841 |
| PSI & PLE (θ) | 0.302 | 0.182 | 0.042 | 0.137 | 0.160 | 0.172 | 0.302 | 0.832 | 0.137 | 0.177 | 0.137 | 0.197 | 0.160 | 0.137 |
| | 0.300 | 0.506 | 0.130 | 0.055 | 0.003 | 0.006 | 0.003 | 0.000 | 0.000 | 0.006 | 0.011 | 0.009 | 0.075 | 0.003 |
| PSI & PLE (α) | 0.126 | 0.194 | 0.126 | 0.137 | 0.126 | 0.435 | 0.392 | 0.562 | 0.573 | 0.490 | 0.734 | 0.284 | 0.226 | 0.126 |
| | 0.820 | 0.209 | 0.000 | 0.003 | 0.032 | 0.014 | 0.364 | 0.048 | 0.112 | 0.048 | 0.026 | 0.003 | 0.000 | 0.000 |
| PSI & PLE (β) | 0.081 | 0.743 | 0.185 | 0.005 | 0.098 | 0.128 | 0.157 | 0.847 | 0.042 | 0.128 | 0.743 | 0.303 | 0.005 | 0.000 |
| | 0.449 | 0.115 | 0.099 | 0.051 | 0.000 | 0.009 | 0.000 | 0.014 | 0.011 | 0.099 | 0.592 | 0.502 | 0.099 | 0.099 |
| PSI & PLI (δ) | 0.827 | 0.697 | 0.341 | 0.254 | 0.224 | 0.014 | 0.246 | 0.711 | 0.224 | 0.224 | 0.324 | 0.675 | 0.711 | 0.827 |
| | 0.372 | 0.795 | 0.372 | 0.669 | 0.382 | 0.795 | 0.620 | 0.372 | 0.372 | 0.372 | 0.382 | 0.620 | 0.562 | 0.971 |
| PSI & PLI (θ) | 0.158 | 0.592 | 0.835 | 0.956 | 0.835 | 0.592 | 0.592 | 0.993 | 0.592 | 0.592 | 0.140 | 0.277 | 0.014 | 0.014 |
| | 0.976 | 0.976 | 0.749 | 0.976 | 0.976 | 0.976 | 0.976 | 0.976 | 0.976 | 0.976 | 0.976 | 0.749 | 0.976 | 0.976 |
| PSI & PLI (α) | 0.424 | 0.622 | 0.424 | 0.903 | 0.506 | 0.878 | 0.861 | 0.808 | 0.770 | 0.662 | 0.820 | 0.506 | 0.210 | 0.000 |
| | 0.850 | 0.436 | 0.088 | 0.436 | 0.552 | 0.552 | 0.723 | 0.436 | 0.641 | 0.194 | 0.106 | 0.088 | 0.000 | 0.088 |
| PSI & PLI (β) | 0.655 | 0.496 | 0.830 | 0.028 | 0.210 | 0.275 | 0.655 | 0.898 | 0.338 | 0.946 | 0.655 | 0.338 | 0.326 | 0.655 |
| | 0.352 | 0.352 | 0.536 | 0.352 | 0.352 | 0.438 | 0.352 | 0.352 | 0.352 | 0.400 | 0.615 | 0.568 | 0.352 | 0.352 |
| PSI & power (δ) | 0.103 | 0.070 | 0.103 | 0.070 | 0.000 | 0.000 | 0.000 | 0.103 | 0.155 | 0.163 | 0.792 | 0.103 | 0.879 | 0.532 |
| | 0.112 | 0.539 | 0.606 | 0.976 | 0.792 | 0.970 | 0.606 | 0.672 | 0.606 | 0.606 | 0.329 | 0.513 | 0.970 | 0.976 |
| PSI & power (θ) | 0.322 | 0.056 | 0.322 | 0.468 | 0.000 | 0.019 | 0.000 | 0.468 | 0.975 | 0.975 | 0.975 | 0.244 | 0.428 | 0.322 |
| | 0.955 | 0.955 | 0.955 | 0.955 | 0.955 | 0.955 | 0.955 | 0.955 | 0.955 | 0.955 | 0.955 | 0.955 | 0.955 | 0.955 |
| PSI & power (α) | 0.742 | 0.084 | 0.911 | 0.911 | 0.084 | 0.532 | 0.084 | 0.614 | 0.867 | 0.614 | 0.532 | 0.911 | 0.859 | 0.932 |
| | 0.648 | 0.648 | 0.648 | 0.764 | 0.764 | 0.648 | 0.648 | 0.648 | 0.648 | 0.648 | 0.764 | 0.769 | 0.648 | 0.648 |
| PSI & power (β) | 0.294 | 0.462 | 0.294 | 0.570 | 0.551 | 0.714 | 0.294 | 0.570 | 0.585 | 0.460 | 0.294 | 0.460 | 0.714 | 0.570 |
| | 0.251 | 0.508 | 0.127 | 0.104 | 0.004 | 0.004 | 0.006 | 0.004 | 0.004 | 0.016 | 0.039 | 0.020 | 0.340 | 0.267 |

C. Correlation coefficients (PSI at PACU) (Pfol N=23; Rem N=24)

| Measures / States | Baseline | | After Loss of consciousness | | | | | Before Recovery of consciousness | | | | | Emergence | |
|---|---|---|---|---|---|---|---|---|---|---|---|---|---|---|
| | EC | EO | LOC1 | LOC2 | LOC3 | LOC4 | LOC5 | ROC1 | ROC2 | ROC3 | ROC4 | ROC5 | EC | EO |
| PSI (PACU) & PLE (δ) | -.15 | .08 | .10 | .04 | .19 | -.09 | -.14 | -.03 | -.10 | -.16 | -.23 | -.16 | -.09 | .09 |
| | -.17 | -.34 | -.09 | -.21 | -.21 | -.07 | -.05 | -.05 | -.04 | -.15 | -.08 | -.08 | -.09 | -.15 |
| PSI (PACU) & PLE (θ) | -.19 | .23 | .05 | -.12 | -.09 | -.20 | -.26 | -.02 | -.17 | -.10 | -.22 | -.13 | .40 | .36 |

| | | | | | | | | | | | | | | |
|---|---|---|---|---|---|---|---|---|---|---|---|---|---|---|
| | .34 | -.27 | .16 | .32 | .41 | .41 | .31 | .25 | .22 | .17 | .15 | .15 | .44 | .58 |
| PSI (PACU) & PLE (α) | .29 | .21 | .16 | .27 | .28 | .28 | .36 | -.04 | .23 | .19 | .13 | -.13 | .33 | .42 |
| | .24 | .08 | .47 | .50 | .36 | .26 | .29 | .49 | .42 | .51 | .52 | .43 | .72 | .70 |
| PSI (PACU) & PLE (β) | -.10 | .02 | -.19 | -.13 | -.22 | -.15 | .08 | -.06 | -.16 | -.17 | .00 | -.02 | .55 | .73 |
| | .43 | .15 | -.26 | -.19 | -.16 | -.16 | -.20 | -.23 | -.20 | -.07 | -.06 | .04 | .36 | .28 |
| PSI (PACU) & PLI (δ) | .09 | .01 | .29 | .22 | -.06 | .14 | .02 | -.13 | .19 | .16 | -.01 | .31 | .17 | -.19 |
| | -.23 | -.02 | -.06 | .11 | .09 | .12 | -.07 | .04 | -.17 | -.27 | -.13 | .06 | .10 | .07 |
| PSI (PACU) & PLI (θ) | .24 | .10 | .16 | .28 | .10 | -.05 | .04 | .12 | .13 | .17 | .45 | .21 | -.61 | -.59 |
| | .42 | -.04 | -.15 | -.02 | -.25 | -.26 | -.32 | .12 | -.20 | -.27 | .12 | -.31 | -.19 | -.30 |
| PSI (PACU) & PLI (α) | -.46 | .16 | -.02 | -.13 | -.13 | -.17 | -.30 | .02 | -.05 | -.03 | .00 | .20 | -.43 | -.68 |
| | .16 | .01 | -.26 | -.26 | -.21 | -.15 | -.12 | -.26 | -.25 | -.43 | -.40 | -.35 | -.63 | -.50 |
| PSI (PACU) & PLI (β) | -.02 | .05 | .19 | .03 | .09 | .10 | -.22 | .11 | .25 | .14 | .13 | .12 | -.24 | -.19 |
| | .05 | .22 | .06 | .06 | .08 | .03 | -.01 | .03 | .14 | -.09 | -.08 | -.07 | -.29 | .19 |
| PSI (PACU) & power (δ) | .32 | .11 | -.03 | -.05 | -.03 | .00 | .05 | .06 | .18 | .05 | .21 | .21 | -.06 | -.19 |
| | -.22 | .24 | -.28 | -.36 | -.34 | -.33 | -.22 | -.21 | -.24 | -.22 | -.26 | -.16 | -.10 | .03 |
| PSI (PACU) & power (θ) | .33 | .12 | .03 | .03 | .03 | -.01 | .05 | .03 | .12 | -.02 | .03 | .13 | -.26 | -.31 |
| | -.43 | .20 | -.39 | -.58 | -.55 | -.54 | -.45 | -.50 | -.48 | -.51 | -.47 | -.45 | -.29 | -.29 |
| PSI (PACU) & power (α) | .12 | .11 | .02 | -.02 | -.01 | -.08 | -.10 | .05 | .04 | -.02 | -.03 | .08 | -.12 | -.09 |
| | -.41 | -.10 | -.43 | -.48 | -.44 | -.34 | -.32 | -.44 | -.44 | -.47 | -.47 | -.40 | -.32 | -.27 |
| PSI (PACU) & power (β) | .13 | .30 | .06 | .05 | .11 | -.04 | -.07 | .03 | .05 | .02 | -.02 | .03 | -.03 | .06 |
| | -.36 | -.12 | -.06 | -.13 | -.10 | -.03 | -.05 | -.04 | -.05 | -.05 | -.03 | .00 | .19 | .26 |
| PSI (PACU) & Time to Aldrete | -- | -- | -- | -- | -- | -- | -- | -- | -- | -- | -- | -- | -.14 | |
| | -- | -- | -- | -- | -- | -- | -- | -- | -- | -- | -- | -- | -.45 | |
| PSI (PACU) & ANI | .46 | .53 | -.18 | .21 | -.20 | .29 | .18 | -.08 | .19 | -.16 | .38 | -.25 | .02 | .20 |
| | .23 | .15 | -.17 | -.37 | -.22 | -.28 | -.24 | -.17 | -.35 | -.26 | -.15 | .00 | .16 | .06 |

D. FDR-corrected *P*-values (PSI at PACU) (Pfol N=23; Rem N=24)

| Measures / States | Baseline | | After Loss of consciousness | | | | | Before Recovery of consciousness | | | | | Emergence | |
|---|---|---|---|---|---|---|---|---|---|---|---|---|---|---|
| | EC | EO | LOC1 | LOC2 | LOC3 | LOC4 | LOC5 | ROC1 | ROC2 | ROC3 | ROC4 | ROC5 | EC | EO |
| PSI (PACU) & PLE (δ) | 0.846 | 0.846 | 0.846 | 0.906 | 0.846 | 0.846 | 0.846 | 0.906 | 0.846 | 0.846 | 0.846 | 0.846 | 0.846 | 0.846 |
| | 0.844 | 0.844 | 0.844 | 0.844 | 0.844 | 0.844 | 0.844 | 0.844 | 0.844 | 0.844 | 0.844 | 0.844 | 0.844 | 0.844 |
| PSI (PACU) & PLE (θ) | 0.779 | 0.779 | 0.885 | 0.804 | 0.804 | 0.779 | 0.779 | 0.935 | 0.779 | 0.804 | 0.779 | 0.804 | 0.622 | 0.622 |
| | 0.272 | 0.346 | 0.498 | 0.272 | 0.165 | 0.165 | 0.272 | 0.386 | 0.429 | 0.498 | 0.498 | 0.498 | 0.165 | 0.045 |
| PSI (PACU) & PLE (α) | 0.432 | 0.517 | 0.585 | 0.432 | 0.432 | 0.432 | 0.432 | 0.860 | 0.517 | 0.539 | 0.604 | 0.604 | 0.432 | 0.432 |
| | 0.289 | 0.720 | 0.039 | 0.035 | 0.112 | 0.265 | 0.212 | 0.035 | 0.060 | 0.035 | 0.035 | 0.060 | 0.001 | 0.001 |
| PSI (PACU) & PLE (β) | 0.987 | 0.987 | 0.987 | 0.987 | 0.987 | 0.987 | 0.987 | 0.987 | 0.987 | 0.987 | 0.987 | 0.987 | 0.044 | 0.001 |

| | | | | | | | | | | | | | | |
|---|---|---|---|---|---|---|---|---|---|---|---|---|---|---|
| | 0.508 | 0.614 | 0.614 | 0.614 | 0.614 | 0.614 | 0.614 | 0.614 | 0.614 | 0.844 | 0.844 | 0.844 | 0.583 | 0.614 |
| PSI (PACU) & PLI (δ) | 0.935 | 0.962 | 0.864 | 0.864 | 0.962 | 0.864 | 0.962 | 0.864 | 0.864 | 0.864 | 0.962 | 0.864 | 0.864 | 0.864 |
| | 0.918 | 0.926 | 0.918 | 0.918 | 0.918 | 0.918 | 0.918 | 0.918 | 0.918 | 0.918 | 0.918 | 0.918 | 0.918 | 0.918 |
| PSI (PACU) & PLI (θ) | 0.733 | 0.770 | 0.770 | 0.673 | 0.770 | 0.866 | 0.872 | 0.770 | 0.770 | 0.770 | 0.140 | 0.770 | 0.023 | 0.023 |
| | 0.489 | 0.929 | 0.663 | 0.929 | 0.489 | 0.489 | 0.489 | 0.678 | 0.588 | 0.489 | 0.678 | 0.489 | 0.588 | 0.489 |
| PSI (PACU) & PLI (α) | 0.184 | 0.873 | 0.986 | 0.873 | 0.873 | 0.873 | 0.572 | 0.986 | 0.986 | 0.986 | 0.986 | 0.873 | 0.197 | 0.004 |
| | 0.563 | 0.981 | 0.367 | 0.367 | 0.452 | 0.575 | 0.619 | 0.367 | 0.367 | 0.160 | 0.177 | 0.255 | 0.013 | 0.084 |
| PSI (PACU) & PLI (β) | 0.935 | 0.935 | 0.874 | 0.935 | 0.874 | 0.874 | 0.874 | 0.874 | 0.874 | 0.874 | 0.874 | 0.874 | 0.874 | 0.874 |
| | 0.958 | 0.958 | 0.958 | 0.958 | 0.958 | 0.958 | 0.979 | 0.958 | 0.958 | 0.958 | 0.958 | 0.958 | 0.958 | 0.958 |
| PSI (PACU) & power (δ) | 0.956 | 0.956 | 0.956 | 0.956 | 0.956 | 0.997 | 0.956 | 0.956 | 0.956 | 0.956 | 0.956 | 0.956 | 0.956 | 0.956 |
| | 0.424 | 0.424 | 0.424 | 0.424 | 0.424 | 0.424 | 0.424 | 0.428 | 0.424 | 0.424 | 0.424 | 0.534 | 0.691 | 0.875 |
| PSI (PACU) & power (θ) | 0.972 | 0.972 | 0.972 | 0.972 | 0.972 | 0.972 | 0.972 | 0.972 | 0.972 | 0.972 | 0.972 | 0.972 | 0.972 | 0.972 |
| | 0.051 | 0.359 | 0.073 | 0.031 | 0.031 | 0.031 | 0.043 | 0.036 | 0.039 | 0.036 | 0.039 | 0.043 | 0.189 | 0.189 |
| PSI (PACU) & power (α) | 0.980 | 0.980 | 0.980 | 0.980 | 0.980 | 0.980 | 0.980 | 0.980 | 0.980 | 0.980 | 0.980 | 0.980 | 0.980 | 0.980 |
| | 0.078 | 0.654 | 0.072 | 0.072 | 0.072 | 0.145 | 0.154 | 0.072 | 0.072 | 0.072 | 0.072 | 0.086 | 0.154 | 0.213 |
| PSI (PACU) & power (β) | 0.942 | 0.942 | 0.942 | 0.942 | 0.942 | 0.942 | 0.942 | 0.942 | 0.942 | 0.942 | 0.942 | 0.942 | 0.942 | 0.942 |
| | 0.957 | 0.957 | 0.957 | 0.957 | 0.957 | 0.957 | 0.957 | 0.957 | 0.957 | 0.957 | 0.957 | 0.982 | 0.957 | 0.957 |
| PSI (PACU) & Time to Aldrete | -- | -- | -- | -- | -- | -- | -- | -- | -- | -- | -- | -- | 0.768 | |
| | -- | -- | -- | -- | -- | -- | -- | -- | -- | -- | -- | -- | 0.071 | |
| PSI (PACU) & ANI | 0.454 | 0.454 | 0.557 | 0.557 | 0.557 | 0.557 | 0.557 | 0.816 | 0.557 | 0.557 | 0.454 | 0.557 | 0.945 | 0.557 |
| | 0.671 | 0.684 | 0.679 | 0.671 | 0.671 | 0.671 | 0.671 | 0.679 | 0.671 | 0.671 | 0.679 | 0.998 | 0.679 | 0.858 |

**Table S1.** Pearson's correlation between PSI and results values.

Pearson's correlation coefficients and Benjamini-Hochberg (FDR) adjusted *P*-values of corresponding coefficients, whereby **(A)** and **(B)** include correlation between PSI values and results values (connectivity and powers at four frequency bands), **(C)** and **(D)** include correlation between PSI at PACU (averaged values of EC and EO emergence) and results values. The blue and red values represent the correlation values in propofol and remimazolam groups, respectively. The highlights colored with 3-steps correspond to the significance level of the FDR-corrected *P*-value (yellow: $P < 0.05$, orange: $P < 0.01$, dark orange: $P < 0.0001$).

A. Correlation coefficients (N=48)

| Measures \ States | Baseline | | After Loss of consciousness | | | | | Before Recovery of consciousness | | | | | Emergence | |
|---|---|---|---|---|---|---|---|---|---|---|---|---|---|---|
| | EC | EO | LOC1 | LOC2 | LOC3 | LOC4 | LOC5 | ROC1 | ROC2 | ROC3 | ROC4 | ROC5 | EC | EO |
| Time to Aldrete & PLE (δ) | .24<br>-.06 | .20<br>.38 | .46<br>-.03 | .41<br>-.03 | .30<br>-.07 | .37<br>-.12 | .43<br>-.14 | .02<br>-.08 | -.07<br>-.04 | -.19<br>.07 | .07<br>-.02 | .30<br>-.03 | -.12<br>.16 | .46<br>.25 |
| Time to Aldrete & PLE (θ) | .33<br>-.32 | .16<br>.20 | .11<br>.04 | .14<br>-.04 | .06<br>-.18 | .09<br>-.06 | -.03<br>-.04 | -.07<br>.01 | -.02<br>.05 | -.22<br>.17 | -.26<br>.19 | -.08<br>.19 | -.12<br>.20 | .24<br>.20 |
| Time to Aldrete & PLE (α) | .41<br>.11 | -.09<br>.17 | .15<br>-.30 | .19<br>-.30 | .15<br>-.24 | .20<br>-.30 | .27<br>-.28 | -.09<br>-.35 | .02<br>-.35 | .09<br>-.34 | -.03<br>-.38 | -.12<br>-.31 | -.04<br>-.45 | .21<br>-.02 |
| Time to Aldrete & PLE (β) | .09<br>-.23 | -.14<br>.22 | -.13<br>.11 | -.31<br>.01 | -.16<br>.08 | -.17<br>.06 | -.24<br>.12 | .05<br>.00 | .10<br>.01 | .21<br>-.16 | .02<br>-.17 | -.10<br>-.23 | .19<br>-.18 | .15<br>.13 |
| Time to Aldrete & PLI (δ) | -.27<br>.13 | -.10<br>-.13 | -.23<br>.11 | -.03<br>.50 | -.32<br>.31 | -.20<br>.07 | -.18<br>.28 | .05<br>.17 | .35<br>-.19 | .23<br>.32 | -.15<br>.19 | -.05<br>.06 | .15<br>-.12 | -.36<br>-.12 |
| Time to Aldrete & PLI (θ) | -.03<br>-.37 | .03<br>-.19 | -.03<br>.32 | -.07<br>.03 | -.27<br>.34 | -.38<br>.20 | -.10<br>-.08 | -.06<br>-.04 | -.16<br>-.13 | .25<br>-.14 | .10<br>-.08 | .04<br>-.04 | .03<br>-.06 | -.21<br>.14 |
| Time to Aldrete & PLI (α) | -.19<br>-.49 | .09<br>-.24 | -.06<br>.30 | -.18<br>.26 | -.11<br>.27 | -.19<br>.37 | -.30<br>.43 | .18<br>.47 | .01<br>.46 | -.08<br>.49 | .01<br>.49 | .03<br>.52 | .09<br>.53 | -.05<br>.23 |
| Time to Aldrete & PLI (β) | .03<br>.17 | .00<br>-.07 | .35<br>.18 | .27<br>.11 | .15<br>.18 | .07<br>.28 | .05<br>.30 | .18<br>.27 | .02<br>.19 | -.14<br>.28 | .04<br>.24 | .18<br>.37 | .41<br>.37 | .08<br>.14 |
| Time to Aldrete & power (δ) | .01<br>.29 | -.03<br>.26 | -.35<br>-.16 | -.19<br>-.18 | -.06<br>-.28 | -.08<br>-.31 | -.04<br>-.30 | -.19<br>-.23 | -.12<br>-.25 | -.10<br>-.15 | -.01<br>-.20 | -.12<br>-.06 | -.18<br>-.11 | .10<br>-.05 |
| Time to Aldrete & power (θ) | -.03<br>.21 | -.06<br>.33 | -.40<br>-.22 | -.16<br>-.25 | -.11<br>-.29 | -.05<br>-.27 | -.08<br>-.29 | -.20<br>-.34 | -.03<br>-.38 | -.03<br>-.23 | .03<br>-.28 | -.01<br>-.16 | -.22<br>-.06 | -.02<br>.09 |
| Time to Aldrete & power (α) | .00<br>.13 | -.03<br>.24 | -.22<br>-.23 | -.10<br>-.17 | -.01<br>-.17 | -.01<br>-.09 | -.09<br>-.13 | -.15<br>-.23 | -.12<br>-.26 | -.05<br>-.20 | .01<br>-.23 | .01<br>-.12 | .09<br>-.06 | .16<br>.02 |
| Time to Aldrete & power (β) | -.01<br>.25 | -.11<br>.21 | -.09<br>.09 | .02<br>.07 | .04<br>.15 | .04<br>.15 | -.14<br>.10 | -.19<br>.03 | -.17<br>.01 | -.14<br>-.04 | -.12<br>-.04 | -.10<br>.05 | -.08<br>.22 | .00<br>.21 |
| Time to Aldrete & PSI | -.02<br>-.06 | .26<br>.07 | .09<br>-.20 | .25<br>-.30 | .29<br>-.37 | .32<br>-.26 | .32<br>-.24 | -.25<br>-.39 | -.17<br>-.41 | -.33<br>-.41 | -.13<br>-.44 | -.21<br>-.32 | .05<br>-.59 | .14<br>-.22 |
| Time to Aldrete & ANI | -.30<br>.28 | .19<br>.30 | .64<br>.06 | .48<br>.14 | .69<br>.10 | .67<br>.24 | .51<br>.20 | .71<br>.07 | .87<br>.17 | .77<br>.19 | .63<br>.29 | .72<br>.13 | .06<br>-.38 | .40<br>-.05 |

B. Benjamini-Hochberg (FDR) corrected *P*-values (N=48)

| Measures / States | Baseline EC | Baseline EO | After Loss of consciousness LOC1 | LOC2 | LOC3 | LOC4 | LOC5 | Before Recovery of consciousness ROC1 | ROC2 | ROC3 | ROC4 | ROC5 | Emergence EC | Emergence EO |
|---|---|---|---|---|---|---|---|---|---|---|---|---|---|---|
| Time to Aldrete & PLE (δ) | 0.460 | 0.522 | 0.168 | 0.175 | 0.314 | 0.218 | 0.168 | 0.941 | 0.802 | 0.522 | 0.802 | 0.314 | 0.717 | 0.168 |
| | 0.911 | 0.911 | 0.911 | 0.911 | 0.911 | 0.911 | 0.911 | 0.911 | 0.911 | 0.911 | 0.911 | 0.911 | 0.911 | 0.911 |
| Time to Aldrete & PLE (θ) | 0.917 | 0.917 | 0.917 | 0.917 | 0.917 | 0.917 | 0.942 | 0.917 | 0.942 | 0.917 | 0.917 | 0.917 | 0.917 | 0.917 |
| | 0.728 | 0.728 | 0.937 | 0.937 | 0.728 | 0.937 | 0.937 | 0.951 | 0.937 | 0.728 | 0.728 | 0.728 | 0.728 | 0.728 |
| Time to Aldrete & PLE (α) | 0.644 | 0.878 | 0.878 | 0.878 | 0.878 | 0.878 | 0.878 | 0.878 | 0.932 | 0.878 | 0.932 | 0.878 | 0.932 | 0.878 |
| | 0.669 | 0.501 | 0.244 | 0.244 | 0.339 | 0.244 | 0.251 | 0.244 | 0.244 | 0.244 | 0.244 | 0.244 | 0.244 | 0.914 |
| Time to Aldrete & PLE (β) | 0.792 | 0.792 | 0.792 | 0.792 | 0.792 | 0.792 | 0.792 | 0.881 | 0.792 | 0.792 | 0.913 | 0.792 | 0.792 | 0.792 |
| | 0.954 | 0.954 | 0.954 | 0.995 | 0.985 | 0.985 | 0.954 | 0.995 | 0.995 | 0.954 | 0.954 | 0.954 | 0.954 | 0.954 |
| Time to Aldrete & PLI (δ) | 0.644 | 0.803 | 0.644 | 0.889 | 0.593 | 0.669 | 0.669 | 0.886 | 0.593 | 0.644 | 0.669 | 0.886 | 0.669 | 0.593 |
| | 0.718 | 0.718 | 0.718 | 0.182 | 0.634 | 0.782 | 0.634 | 0.718 | 0.718 | 0.634 | 0.718 | 0.782 | 0.718 | 0.718 |
| Time to Aldrete & PLI (θ) | 0.902 | 0.902 | 0.902 | 0.902 | 0.902 | 0.902 | 0.902 | 0.902 | 0.902 | 0.902 | 0.902 | 0.902 | 0.902 | 0.902 |
| | 0.602 | 0.887 | 0.602 | 0.887 | 0.602 | 0.887 | 0.887 | 0.887 | 0.887 | 0.887 | 0.887 | 0.887 | 0.887 | 0.887 |
| Time to Aldrete & PLI (α) | 0.979 | 0.979 | 0.979 | 0.979 | 0.979 | 0.979 | 0.979 | 0.979 | 0.979 | 0.979 | 0.979 | 0.979 | 0.979 | 0.979 |
| | 0.045 | 0.271 | 0.209 | 0.254 | 0.254 | 0.121 | 0.060 | 0.046 | 0.046 | 0.045 | 0.045 | 0.045 | 0.045 | 0.271 |
| Time to Aldrete & PLI (β) | 0.993 | 0.993 | 0.644 | 0.952 | 0.993 | 0.993 | 0.993 | 0.993 | 0.993 | 0.993 | 0.993 | 0.993 | 0.644 | 0.993 |
| | 0.547 | 0.744 | 0.547 | 0.648 | 0.547 | 0.457 | 0.457 | 0.457 | 0.547 | 0.457 | 0.516 | 0.457 | 0.457 | 0.614 |
| Time to Aldrete & power (δ) | 0.979 | 0.979 | 0.979 | 0.979 | 0.979 | 0.979 | 0.979 | 0.979 | 0.979 | 0.979 | 0.979 | 0.979 | 0.979 | 0.979 |
| | 0.548 | 0.548 | 0.612 | 0.612 | 0.548 | 0.548 | 0.548 | 0.548 | 0.548 | 0.612 | 0.602 | 0.817 | 0.712 | 0.817 |
| Time to Aldrete & power (θ) | 0.963 | 0.963 | 0.728 | 0.963 | 0.963 | 0.963 | 0.963 | 0.963 | 0.963 | 0.963 | 0.963 | 0.963 | 0.963 | 0.963 |
| | 0.409 | 0.409 | 0.409 | 0.409 | 0.409 | 0.409 | 0.409 | 0.409 | 0.409 | 0.409 | 0.409 | 0.544 | 0.769 | 0.743 |
| Time to Aldrete & power (α) | 0.992 | 0.992 | 0.992 | 0.992 | 0.992 | 0.992 | 0.992 | 0.992 | 0.992 | 0.992 | 0.992 | 0.992 | 0.992 | 0.992 |
| | 0.731 | 0.731 | 0.731 | 0.731 | 0.731 | 0.781 | 0.731 | 0.731 | 0.731 | 0.731 | 0.731 | 0.731 | 0.827 | 0.911 |
| Time to Aldrete & power (β) | 0.996 | 0.996 | 0.996 | 0.996 | 0.996 | 0.996 | 0.996 | 0.996 | 0.996 | 0.996 | 0.996 | 0.996 | 0.996 | 0.996 |
| | 0.970 | 0.970 | 0.970 | 0.970 | 0.970 | 0.970 | 0.970 | 0.970 | 0.974 | 0.970 | 0.970 | 0.970 | 0.970 | 0.970 |
| Time to Aldrete & PSI | 0.913 | 0.488 | 0.789 | 0.488 | 0.488 | 0.488 | 0.488 | 0.488 | 0.672 | 0.488 | 0.703 | 0.558 | 0.892 | 0.703 |
| | 0.801 | 0.801 | 0.394 | 0.263 | 0.180 | 0.330 | 0.365 | 0.162 | 0.161 | 0.161 | 0.161 | 0.244 | 0.028 | 0.387 |
| Time to Aldrete & ANI | 0.727 | 0.870 | 0.221 | 0.378 | 0.221 | 0.221 | 0.378 | 0.221 | 0.143 | 0.221 | 0.221 | 0.221 | 0.913 | 0.482 |
| | 0.861 | 0.861 | 0.861 | 0.861 | 0.861 | 0.861 | 0.861 | 0.861 | 0.861 | 0.861 | 0.861 | 0.861 | 0.861 | 0.861 |

C. Correlation coefficients (zero excluded) (Pfol *N*=7; Rem *N*=17)

| Measures \ States | Baseline | | After Loss of consciousness | | | | | Before Recovery of consciousness | | | | | Emergence | |
|---|---|---|---|---|---|---|---|---|---|---|---|---|---|---|
| | EC | EO | LOC1 | LOC2 | LOC3 | LOC4 | LOC5 | ROC1 | ROC2 | ROC3 | ROC4 | ROC5 | EC | EO |
| Time to Aldrete & PLE (δ) | .29 | .47 | .06 | -.05 | -.13 | -.05 | .22 | -.13 | -.23 | -.51 | -.17 | .25 | -.40 | .72 |
| | -.13 | .24 | .15 | .07 | .11 | .03 | .15 | .15 | .18 | .24 | .28 | .20 | .22 | .36 |
| Time to Aldrete & PLE (θ) | .46 | -.20 | -.01 | -.18 | -.16 | .21 | .02 | -.12 | -.20 | -.03 | -.13 | -.20 | -.52 | .67 |
| | -.57 | .08 | -.15 | -.22 | -.28 | -.22 | -.19 | -.24 | -.15 | -.10 | .02 | .05 | .06 | -.11 |
| Time to Aldrete & PLE (α) | .39 | .39 | .39 | .39 | .59 | .59 | .63 | .63 | .63 | .63 | .63 | .63 | .65 | .66 |
| | .12 | .06 | -.53 | -.50 | -.47 | -.47 | -.60 | -.78 | -.77 | -.75 | -.68 | -.53 | -.52 | -.25 |
| Time to Aldrete & PLE (β) | .63 | -.34 | -.43 | -.36 | -.24 | -.47 | -.91 | -.56 | -.30 | .66 | .11 | -.26 | .45 | .10 |
| | -.15 | .06 | .26 | .19 | .29 | .29 | .34 | .42 | .38 | .31 | .29 | .20 | -.12 | -.07 |
| Time to Aldrete & PLI (δ) | -.28 | .16 | .22 | .17 | -.08 | -.05 | -.10 | .89 | .41 | .42 | .64 | -.05 | .19 | -.70 |
| | -.03 | -.15 | -.23 | .23 | .00 | .05 | .00 | -.14 | -.49 | .12 | -.21 | .06 | .03 | -.28 |
| Time to Aldrete & PLI (θ) | .52 | .73 | -.29 | .07 | -.60 | -.62 | -.33 | -.30 | .11 | .01 | -.22 | -.07 | .63 | -.19 |
| | -.59 | -.18 | .36 | .23 | .30 | .32 | .22 | .21 | .22 | .00 | -.15 | -.02 | .05 | .33 |
| Time to Aldrete & PLI (α) | -.21 | .81 | .22 | -.18 | .07 | -.30 | -.55 | .13 | .08 | .22 | .29 | .11 | .21 | .03 |
| | -.54 | .02 | .25 | .36 | .24 | .40 | .42 | .66 | .62 | .69 | .60 | .46 | .53 | .38 |
| Time to Aldrete & PLI (β) | -.39 | .36 | .43 | .29 | .12 | .15 | .30 | .65 | .56 | -.05 | .28 | .25 | .51 | -.10 |
| | .15 | -.35 | .06 | .06 | .05 | .11 | .09 | .16 | .04 | .17 | .07 | .20 | .23 | .21 |
| Time to Aldrete & power (δ) | -.32 | -.13 | .23 | .21 | .25 | .16 | .08 | .60 | .40 | .43 | .48 | .27 | .81 | -.78 |
| | .27 | -.25 | -.02 | .05 | .07 | .10 | .05 | .20 | .21 | .07 | .07 | .10 | -.01 | .01 |
| Time to Aldrete & power (θ) | -.02 | .17 | .16 | .19 | .16 | .04 | .04 | .32 | .13 | .04 | .37 | .60 | .95 | .24 |
| | .40 | -.21 | .14 | .19 | .18 | .16 | .19 | .40 | .48 | .43 | .29 | .32 | .11 | .20 |
| Time to Aldrete & power (α) | .13 | .26 | .19 | .17 | .17 | .08 | .07 | .00 | -.02 | -.08 | .25 | .43 | .49 | .30 |
| | .12 | -.19 | .16 | .14 | .12 | .11 | .17 | .34 | .38 | .41 | .41 | .39 | .33 | .26 |
| Time to Aldrete & power (β) | .22 | .08 | -.20 | -.07 | .00 | .08 | .35 | -.20 | -.39 | -.28 | .08 | .03 | .14 | .61 |
| | .09 | -.10 | -.12 | -.12 | -.16 | -.15 | -.11 | -.14 | -.14 | -.12 | -.11 | -.07 | .00 | .05 |
| Time to Aldrete & PSI | .14 | .02 | -.10 | -.09 | -.02 | .03 | .09 | -.43 | -.41 | -.72 | -.33 | -.26 | -.29 | .09 |
| | .38 | .14 | -.10 | .03 | .13 | .09 | -.06 | .11 | .21 | .22 | .23 | .26 | .23 | .22 |
| Time to Aldrete & ANI | -.30 | .19 | .64 | .48 | .69 | .67 | .51 | .71 | .87 | .77 | .63 | .72 | .06 | .40 |
| | .28 | .30 | .06 | .14 | .10 | .24 | .20 | .07 | .17 | .19 | .29 | .13 | -.38 | -.05 |

D. Benjamini-Hochberg (FDR) corrected *P*-values (zero excluded) (Pfol *N*=7; Rem *N*=17)

| Measures / States | Baseline | | After Loss of consciousness | | | | | Before Recovery of consciousness | | | | | Emergence | |
|---|---|---|---|---|---|---|---|---|---|---|---|---|---|---|
| | EC | EO | LOC1 | LOC2 | LOC3 | LOC4 | LOC5 | ROC1 | ROC2 | ROC3 | ROC4 | ROC5 | EC | EO |
| Time to Aldrete & PLE (δ) | 0.913 | 0.913 | 0.913 | 0.913 | 0.913 | 0.913 | 0.913 | 0.913 | 0.913 | 0.913 | 0.913 | 0.913 | 0.913 | 0.910 |
| | 0.799 | 0.799 | 0.799 | 0.859 | 0.802 | 0.920 | 0.799 | 0.799 | 0.799 | 0.799 | 0.799 | 0.799 | 0.799 | 0.799 |
| Time to Aldrete & PLE (θ) | 0.990 | 0.990 | 0.990 | 0.990 | 0.990 | 0.990 | 0.990 | 0.990 | 0.990 | 0.990 | 0.990 | 0.990 | 0.990 | 0.990 |
| | 0.252 | 0.910 | 0.910 | 0.910 | 0.910 | 0.910 | 0.910 | 0.910 | 0.910 | 0.910 | 0.944 | 0.910 | 0.910 | 0.910 |
| Time to Aldrete & PLE (α) | 0.967 | 0.967 | 0.967 | 0.967 | 0.968 | 0.967 | 0.967 | 0.967 | 0.967 | 0.967 | 0.967 | 0.967 | 0.967 | 0.967 |
| | 0.698 | 0.821 | 0.056 | 0.065 | 0.071 | 0.071 | 0.031 | 0.000 | 0.000 | 0.000 | 0.007 | 0.056 | 0.060 | 0.380 |
| Time to Aldrete & PLE (β) | 0.611 | 0.697 | 0.666 | 0.697 | 0.697 | 0.666 | 0.056 | 0.666 | 0.697 | 0.611 | 0.825 | 0.697 | 0.666 | 0.825 |
| | 0.709 | 0.821 | 0.541 | 0.652 | 0.516 | 0.516 | 0.516 | 0.516 | 0.516 | 0.516 | 0.516 | 0.652 | 0.763 | 0.821 |
| Time to Aldrete & PLI (δ) | 0.913 | 0.913 | 0.913 | 0.913 | 0.913 | 0.913 | 0.913 | 0.098 | 0.913 | 0.913 | 0.574 | 0.913 | 0.913 | 0.539 |
| | 0.996 | 0.996 | 0.996 | 0.996 | 0.996 | 0.996 | 0.996 | 0.996 | 0.658 | 0.996 | 0.996 | 0.996 | 0.996 | 0.996 |
| Time to Aldrete & PLI (θ) | 0.636 | 0.553 | 0.931 | 0.951 | 0.553 | 0.553 | 0.931 | 0.931 | 0.951 | 0.989 | 0.951 | 0.951 | 0.553 | 0.951 |
| | 0.182 | 0.701 | 0.641 | 0.641 | 0.641 | 0.641 | 0.641 | 0.641 | 0.641 | 0.986 | 0.708 | 0.986 | 0.986 | 0.641 |
| Time to Aldrete & PLI (α) | 0.949 | 0.364 | 0.949 | 0.949 | 0.949 | 0.949 | 0.949 | 0.949 | 0.949 | 0.949 | 0.949 | 0.949 | 0.949 | 0.949 |
| | 0.068 | 0.951 | 0.387 | 0.204 | 0.387 | 0.166 | 0.165 | 0.028 | 0.037 | 0.028 | 0.042 | 0.132 | 0.068 | 0.190 |
| Time to Aldrete & PLI (β) | 0.823 | 0.823 | 0.823 | 0.823 | 0.900 | 0.900 | 0.823 | 0.823 | 0.823 | 0.913 | 0.823 | 0.823 | 0.823 | 0.900 |
| | 0.864 | 0.864 | 0.864 | 0.864 | 0.864 | 0.864 | 0.864 | 0.864 | 0.864 | 0.864 | 0.864 | 0.864 | 0.864 | 0.864 |
| Time to Aldrete & power (δ) | 0.822 | 0.843 | 0.822 | 0.822 | 0.822 | 0.843 | 0.869 | 0.723 | 0.822 | 0.822 | 0.822 | 0.822 | 0.266 | 0.266 |
| | 0.962 | 0.962 | 0.962 | 0.962 | 0.962 | 0.962 | 0.962 | 0.962 | 0.962 | 0.962 | 0.962 | 0.962 | 0.962 | 0.962 |
| Time to Aldrete & power (θ) | 0.972 | 0.972 | 0.972 | 0.972 | 0.972 | 0.972 | 0.972 | 0.972 | 0.972 | 0.972 | 0.972 | 0.972 | 0.014 | 0.972 |
| | 0.385 | 0.626 | 0.645 | 0.626 | 0.626 | 0.632 | 0.626 | 0.385 | 0.385 | 0.385 | 0.586 | 0.586 | 0.661 | 0.626 |
| Time to Aldrete & power (α) | 0.992 | 0.992 | 0.992 | 0.992 | 0.992 | 0.992 | 0.992 | 0.992 | 0.992 | 0.992 | 0.992 | 0.992 | 0.992 | 0.992 |
| | 0.667 | 0.667 | 0.667 | 0.667 | 0.667 | 0.667 | 0.667 | 0.467 | 0.467 | 0.467 | 0.467 | 0.467 | 0.467 | 0.634 |
| Time to Aldrete & power (β) | 0.992 | 0.992 | 0.992 | 0.992 | 0.992 | 0.992 | 0.992 | 0.992 | 0.992 | 0.992 | 0.992 | 0.992 | 0.992 | 0.992 |
| | 0.920 | 0.920 | 0.920 | 0.920 | 0.920 | 0.920 | 0.920 | 0.920 | 0.920 | 0.920 | 0.920 | 0.929 | 0.993 | 0.929 |
| Time to Aldrete & PSI | 0.965 | 0.965 | 0.965 | 0.965 | 0.965 | 0.965 | 0.965 | 0.965 | 0.965 | 0.965 | 0.965 | 0.965 | 0.965 | 0.965 |
| | 0.832 | 0.851 | 0.851 | 0.923 | 0.851 | 0.851 | 0.890 | 0.851 | 0.832 | 0.832 | 0.832 | 0.832 | 0.832 | 0.832 |
| Time to Aldrete & ANI | 0.727 | 0.870 | 0.221 | 0.378 | 0.221 | 0.221 | 0.378 | 0.221 | 0.143 | 0.221 | 0.221 | 0.221 | 0.913 | 0.482 |
| | 0.861 | 0.861 | 0.861 | 0.861 | 0.861 | 0.861 | 0.861 | 0.861 | 0.861 | 0.861 | 0.861 | 0.861 | 0.861 | 0.861 |

**Table S2.** Pearson's correlation between Time to Aldrete 9 and results values.

Pearson's correlation coefficients between time to Aldrete 9 and measured values of propofol and remimazolam in each state, whereby **(A)** and **(B)** include correlation results for all patients (*N*=48), **(C)** and **(D)** include correlation results for patients except those with zero at 'time to Aldrete 9' (Propofol = 7; remimazolam = 17). **(B)** and **(D)** represent Benjamini-Hochberg (FDR) adjusted *P*-values of corresponding coefficients. The blue and red values represent the correlation values in propofol and remimazolam groups, respectively. The highlights colored with 3-steps correspond to the significance level of the FDR-corrected *P*-value (yellow: $P < 0.05$, orange: $P < 0.01$, dark orange: $P < 0.0001$).

A. ANI & results values

| Measures / States | Baseline | | | | After Loss of consciousness | | | | | | | | | | Before Recovery of consciousness | | | | | | | | | | Emergence | | | |
|---|---|---|---|---|---|---|---|---|---|---|---|---|---|---|---|---|---|---|---|---|---|---|---|---|---|---|---|---|
| | EC | | EO | | LOC1 | | LOC2 | | LOC3 | | LOC4 | | LOC5 | | ROC1 | | ROC2 | | ROC3 | | ROC4 | | ROC5 | | EC | | EO | |
| ANI & PLE (δ) | -.28 | -.10 | .20 | -.31 | -.15 | -.13 | -.10 | .25 | -.65 | -.27 | -.26 | -.35 | -.44 | .02 | -.06 | .04 | -.46 | -.07 | -.47 | .08 | -.68 | -.04 | -.23 | -.20 | -.35 | -.07 | -.13 | -.18 |
| ANI & PLE (θ) | -.20 | .24 | .16 | .03 | .06 | -.07 | .22 | .08 | -.01 | -.42 | .08 | -.38 | .09 | -.03 | -.11 | -.20 | .07 | -.12 | -.11 | .14 | -.43 | -.10 | -.35 | -.37 | -.16 | .21 | -.24 | .27 |
| ANI & PLE (α) | .10 | .37 | -.16 | -.01 | -.16 | -.02 | .13 | -.28 | -.50 | .10 | -.03 | -.03 | .12 | -.10 | -.12 | -.07 | .19 | -.17 | .20 | -.26 | .20 | -.42 | -.20 | -.29 | .05 | .27 | -.01 | .00 |
| ANI & PLE (β) | .12 | .20 | -.11 | -.33 | -.12 | .20 | -.16 | -.05 | .46 | .16 | -.12 | .15 | .06 | .00 | .11 | .00 | .16 | .02 | .11 | -.19 | .44 | .04 | .24 | .34 | -.03 | -.13 | .19 | .36 |
| ANI & PLI (δ) | .49 | .11 | .12 | .03 | .00 | -.03 | .05 | .01 | .55 | .29 | .44 | .24 | .16 | -.25 | .20 | -.15 | .67 | -.15 | .28 | -.23 | .31 | -.31 | .08 | .06 | .37 | -.08 | .35 | -.10 |
| ANI & PLI (θ) | .39 | -.19 | .29 | -.14 | -.15 | -.15 | -.02 | -.29 | -.14 | .03 | -.36 | .24 | -.16 | .18 | .04 | -.42 | -.15 | -.03 | .04 | -.28 | .21 | .02 | .27 | .25 | -.21 | -.11 | .02 | -.42 |
| ANI & PLI (α) | -.13 | -.34 | .27 | -.16 | .30 | -.18 | .25 | .06 | .40 | -.19 | .03 | .15 | -.14 | .27 | .06 | .08 | -.22 | .15 | -.25 | .14 | -.10 | .51 | .30 | .36 | -.39 | -.26 | -.25 | -.11 |
| ANI & PLI (β) | -.11 | .02 | -.21 | -.13 | .23 | -.33 | .18 | .03 | -.52 | -.19 | -.03 | .09 | -.16 | .16 | .14 | .08 | -.11 | -.13 | -.27 | .22 | -.17 | .02 | .17 | -.20 | -.30 | -.14 | -.38 | -.29 |
| ANI & power (δ) | .23 | .08 | .03 | .04 | .29 | .08 | .01 | .10 | .47 | .00 | .34 | -.16 | .42 | -.12 | .27 | -.19 | .30 | -.11 | .07 | -.08 | .27 | -.22 | .20 | -.12 | .03 | -.36 | .24 | .17 |
| ANI & power (θ) | .18 | .02 | .13 | .00 | .22 | -.10 | -.08 | .19 | .21 | -.10 | .14 | -.23 | .26 | -.12 | .53 | -.09 | .19 | -.11 | -.13 | .04 | .01 | -.15 | .18 | -.16 | -.02 | -.42 | .23 | .05 |
| ANI & power (α) | -.06 | -.21 | .25 | .04 | .22 | -.07 | .02 | .28 | .26 | -.23 | .05 | -.40 | .25 | -.10 | .44 | -.09 | .30 | -.03 | -.16 | .24 | -.11 | -.01 | -.04 | -.25 | -.18 | -.31 | -.05 | .28 |
| ANI & power (β) | -.16 | -.04 | .38 | .02 | .18 | -.13 | .13 | .15 | .00 | -.39 | .06 | -.46 | .43 | -.11 | .38 | -.19 | .32 | -.18 | -.10 | .08 | -.09 | -.26 | -.14 | -.44 | -.11 | -.07 | -.13 | .12 |
| ANI & PSI | -.03 | .48 | .12 | -.08 | .03 | -.02 | .23 | -.06 | -.39 | -.36 | -.27 | -.26 | -.22 | -.12 | -.38 | -.06 | -.16 | -.14 | -.34 | -.07 | -.10 | -.23 | .10 | -.27 | -.08 | .21 | .21 | -.03 |
| ANI & Time to Aldrete | -.30 | .28 | .19 | .30 | .64 | .06 | .48 | .14 | .69 | .10 | .67 | .24 | .51 | .20 | .71 | .07 | .87 | .17 | .77 | .19 | .63 | .29 | .72 | .13 | .06 | -.38 | .40 | -.05 |
| Sub no. | P(19)/R(20) | | P(13)/R(16) | | P(19)/R(24) | | P(22)/R(24) | | P(21)/R(22) | | P(22)/R(22) | | P(21)/R(23) | | P(20)/R(22) | | P(22)/R(21) | | P(22)/R(22) | | P(21)/R(21) | | P(20)/R(21) | | P(18)/R(18) | | P(19)/R(19) | |
| Time to Aldrete sub no. | P(5)/R(13) | | P(4)/R(10) | | P(7)/R(17) | | P(7)/R(17) | | P(7)/R(15) | | P(7)/R(15) | | P(7)/R(16) | | P(7)/R(15) | | P(7)/R(14) | | P(7)/R(15) | | P(7)/R(15) | | P(7)/R(14) | | P(6)/R(14) | | P(7)/R(13) | |

B. Benjamini-Hochberg (FDR) corrected *P*-values

| Measures / States | Baseline | | | | After Loss of consciousness | | | | | | | | | | Before Recovery of consciousness | | | | | | | | | | Emergence | | | |
|---|---|---|---|---|---|---|---|---|---|---|---|---|---|---|---|---|---|---|---|---|---|---|---|---|---|---|---|---|
| | EC | | EO | | LOC1 | | LOC2 | | LOC3 | | LOC4 | | LOC5 | | ROC1 | | ROC2 | | ROC3 | | ROC4 | | ROC5 | | EC | | EO | |
| ANI & PLE (δ) | 0.436 | 0.922 | 0.682 | 0.875 | 0.682 | 0.922 | 0.720 | 0.875 | 0.007 | 0.875 | 0.436 | 0.875 | 0.132 | 0.922 | 0.786 | 0.922 | 0.112 | 0.922 | 0.112 | 0.922 | 0.007 | 0.922 | 0.516 | 0.922 | 0.369 | 0.922 | 0.683 | 0.922 |

| | | | | | | | | | | | | | | |
|---|---|---|---|---|---|---|---|---|---|---|---|---|---|---|
| ANI & PLE (θ) | 0.880 / 0.806 | 0.880 / 0.903 | 0.883 / 0.861 | 0.880 / 0.861 | 0.977 / 0.448 | 0.880 / 0.448 | 0.880 / 0.903 | 0.880 / 0.806 | 0.880 / 0.861 | 0.880 / 0.861 | 0.770 / 0.861 | 0.880 / 0.448 | 0.880 / 0.806 | 0.880 / 0.806 |
| ANI & PLE (α) | 0.854 / 0.665 | 0.853 / 0.997 | 0.853 / 0.997 | 0.853 / 0.665 | 0.280 / 0.997 | 0.954 / 0.997 | 0.853 / 0.997 | 0.853 / 0.997 | 0.853 / 0.938 | 0.853 / 0.665 | 0.853 / 0.665 | 0.853 / 0.665 | 0.954 / 0.665 | 0.960 / 0.997 |
| ANI & PLE (β) | 0.812 / 0.884 | 0.854 / 0.884 | 0.812 / 0.884 | 0.812 / 0.987 | 0.315 / 0.884 | 0.812 / 0.884 | 0.856 / 0.987 | 0.812 / 0.987 | 0.812 / 0.987 | 0.812 / 0.884 | 0.315 / 0.987 | 0.812 / 0.884 | 0.905 / 0.949 | 0.812 / 0.884 |
| ANI & PLI (δ) | 0.133 / 0.980 | 0.846 / 0.980 | 0.994 / 0.980 | 0.893 / 0.980 | 0.070 / 0.829 | 0.133 / 0.829 | 0.675 / 0.829 | 0.628 / 0.980 | 0.014 / 0.980 | 0.350 / 0.829 | 0.346 / 0.829 | 0.846 / 0.980 | 0.324 / 0.980 | 0.324 / 0.980 |
| ANI & PLI (θ) | 0.721 / 0.723 | 0.778 / 0.857 | 0.778 / 0.748 | 0.940 / 0.646 | 0.778 / 0.919 | 0.721 / 0.646 | 0.778 / 0.723 | 0.940 / 0.490 | 0.778 / 0.919 | 0.940 / 0.646 | 0.778 / 0.919 | 0.778 / 0.646 | 0.778 / 0.857 | 0.940 / 0.490 |
| ANI & PLI (α) | 0.760 / 0.639 | 0.577 / 0.697 | 0.565 / 0.697 | 0.565 / 0.780 | 0.565 / 0.697 | 0.885 / 0.697 | 0.750 / 0.697 | 0.857 / 0.766 | 0.565 / 0.697 | 0.565 / 0.697 | 0.796 / 0.252 | 0.565 / 0.639 | 0.565 / 0.697 | 0.565 / 0.763 |
| ANI & PLI (β) | 0.713 / 0.934 | 0.682 / 0.900 | 0.682 / 0.900 | 0.682 / 0.934 | 0.224 / 0.900 | 0.901 / 0.900 | 0.682 / 0.900 | 0.700 / 0.900 | 0.713 / 0.900 | 0.682 / 0.900 | 0.682 / 0.934 | 0.682 / 0.900 | 0.682 / 0.900 | 0.682 / 0.900 |
| ANI & power (δ) | 0.549 / 0.873 | 0.975 / 0.963 | 0.518 / 0.873 | 0.975 / 0.873 | 0.420 / 0.984 | 0.518 / 0.873 | 0.420 / 0.873 | 0.518 / 0.873 | 0.518 / 0.873 | 0.962 / 0.873 | 0.518 / 0.873 | 0.563 / 0.873 | 0.975 / 0.873 | 0.549 / 0.873 |
| ANI & power (θ) | 0.795 / 0.996 | 0.837 / 0.996 | 0.795 / 0.970 | 0.837 / 0.970 | 0.795 / 0.970 | 0.806 / 0.970 | 0.795 / 0.970 | 0.224 / 0.970 | 0.795 / 0.970 | 0.806 / 0.996 | 0.973 / 0.970 | 0.795 / 0.970 | 0.973 / 0.970 | 0.795 / 0.996 |
| ANI & power (α) | 0.924 / 0.644 | 0.858 / 0.954 | 0.858 / 0.954 | 0.924 / 0.612 | 0.858 / 0.612 | 0.924 / 0.612 | 0.858 / 0.954 | 0.714 / 0.954 | 0.858 / 0.954 | 0.858 / 0.612 | 0.924 / 0.954 | 0.924 / 0.612 | 0.858 / 0.612 | 0.924 / 0.612 |
| ANI & power (β) | 0.823 / 0.928 | 0.686 / 0.928 | 0.823 / 0.862 | 0.823 / 0.862 | 0.997 / 0.336 | 0.849 / 0.329 | 0.663 / 0.862 | 0.663 / 0.862 | 0.663 / 0.862 | 0.823 / 0.910 | 0.823 / 0.862 | 0.823 / 0.329 | 0.823 / 0.910 | 0.823 / 0.862 |
| ANI & PSI | 0.908 / 0.518 | 0.870 / 0.935 | 0.908 / 0.935 | 0.805 / 0.935 | 0.560 / 0.686 | 0.756 / 0.844 | 0.805 / 0.935 | 0.560 / 0.935 | 0.826 / 0.935 | 0.560 / 0.935 | 0.870 / 0.910 | 0.870 / 0.844 | 0.870 / 0.931 | 0.812 / 0.935 |
| ANI & Time to Aldrete | 0.727 / 0.861 | 0.870 / 0.861 | 0.221 / 0.861 | 0.378 / 0.861 | 0.221 / 0.861 | 0.221 / 0.861 | 0.378 / 0.861 | 0.221 / 0.861 | 0.143 / 0.861 | 0.221 / 0.861 | 0.221 / 0.861 | 0.221 / 0.861 | 0.913 / 0.861 | 0.482 / 0.861 |
| Sub no. | P(19)/R(20) | P(13)/R(16) | P(19)/R(24) | P(22)/R(24) | P(21)/R(22) | P(22)/R(22) | P(21)/R(23) | P(20)/R(22) | P(22)/R(21) | P(22)/R(22) | P(21)/R(21) | P(20)/R(21) | P(18)/R(18) | P(19)/R(19) |
| Time to Aldrete sub no. | P(5)/R(13) | P(4)/R(10) | P(7)/R(17) | P(7)/R(17) | P(7)/R(15) | P(7)/R(15) | P(7)/R(16) | P(7)/R(15) | P(7)/R(14) | P(7)/R(15) | P(7)/R(15) | P(7)/R(14) | P(6)/R(14) | P(7)/R(13) |

C. ANI (PACU) & results values (Pfol N=17; Rem N=14)

| Measures / States | Baseline | | After Loss of consciousness | | | | | Before Recovery of consciousness | | | | | Emergence | |
|---|---|---|---|---|---|---|---|---|---|---|---|---|---|---|
| | EC | EO | LOC1 | LOC2 | LOC3 | LOC4 | LOC5 | ROC1 | ROC2 | ROC3 | ROC4 | ROC5 | EC | EO |
| ANI (PACU) & PLE (δ) | .29 / .08 | .09 / -.24 | -.05 / -.12 | -.39 / -.13 | -.51 / -.19 | -.53 / -.16 | -.52 / -.12 | -.32 / -.25 | -.24 / -.27 | -.40 / -.31 | -.30 / -.42 | -.28 / -.43 | -.45 / -.34 | -.08 / -.27 |
| ANI (PACU) & PLE (θ) | -.23 / .46 | .07 / .11 | .54 / .05 | .08 / .04 | .01 / .19 | .03 / .10 | .05 / .24 | .27 / -.05 | .21 / .06 | .50 / .22 | .39 / .06 | .29 / .14 | -.20 / .04 | -.30 / .19 |
| ANI (PACU) & PLE (α) | -.12 / -.19 | -.21 / .31 | .47 / .32 | .19 / .39 | -.21 / .50 | -.02 / .34 | .09 / .29 | .29 / .24 | .27 / .19 | .16 / -.01 | .35 / -.07 | .37 / -.11 | -.10 / .19 | -.02 / -.19 |
| ANI (PACU) & PLE (β) | .32 | -.10 | -.22 | .13 | .24 | .02 | -.21 | -.27 | -.30 | -.17 | .01 | .14 | .16 | .14 |

| | | | | | | | | | | | | | | |
|---|---|---|---|---|---|---|---|---|---|---|---|---|---|---|
| | -.27 | -.18 | -.36 | -.34 | -.24 | -.12 | -.26 | -.04 | -.05 | .01 | .03 | .20 | .11 | .20 |
| ANI (PACU) & PLI (δ) | -.07 | .32 | .30 | .37 | .36 | .52 | .40 | .39 | .49 | .56 | .27 | .22 | .50 | .33 |
| | .10 | -.14 | -.20 | -.11 | .39 | .06 | -.28 | -.12 | .25 | .14 | .01 | .14 | .21 | -.04 |
| ANI (PACU) & PLI (θ) | -.06 | .06 | -.40 | -.25 | .04 | -.07 | .03 | -.33 | -.04 | -.53 | -.49 | -.61 | -.34 | .05 |
| | .00 | .10 | -.06 | .09 | -.77 | .17 | .01 | -.52 | .00 | -.29 | .20 | .11 | .03 | -.34 |
| ANI (PACU) & PLI (α) | -.01 | .31 | -.38 | -.25 | .09 | -.05 | -.18 | -.34 | -.26 | -.14 | -.27 | -.32 | -.32 | -.25 |
| | .15 | -.29 | -.29 | -.27 | -.46 | -.33 | -.10 | -.04 | -.06 | -.05 | .23 | .31 | -.24 | .13 |
| ANI (PACU) & PLI (β) | -.13 | .22 | -.28 | -.41 | -.47 | -.36 | -.03 | -.15 | .05 | -.23 | -.21 | -.37 | -.51 | -.52 |
| | .16 | -.18 | .17 | .11 | -.15 | -.21 | -.03 | -.16 | -.13 | -.09 | .04 | -.16 | -.47 | -.33 |
| ANI (PACU) & power (δ) | .11 | .02 | .03 | .37 | .61 | .66 | .67 | .22 | .27 | .32 | .30 | .26 | .12 | .10 |
| | .04 | .36 | -.08 | .01 | -.08 | -.05 | -.13 | -.05 | -.04 | .13 | .03 | .55 | -.03 | .29 |
| ANI (PACU) & power (θ) | .34 | .14 | -.14 | .13 | .43 | .44 | .42 | -.06 | .05 | -.02 | .07 | .02 | -.02 | .14 |
| | -.02 | .33 | -.26 | -.12 | -.22 | -.12 | -.26 | -.29 | -.37 | -.25 | -.35 | -.09 | -.25 | .13 |
| ANI (PACU) & power (α) | .28 | .21 | .06 | .22 | .49 | .37 | .38 | .07 | .17 | .20 | .16 | .09 | -.10 | -.11 |
| | .22 | .37 | -.24 | -.14 | -.18 | -.12 | -.13 | -.23 | -.22 | -.09 | -.19 | -.02 | -.26 | .42 |
| ANI (PACU) & power (β) | .08 | .18 | .47 | .27 | .40 | .39 | .46 | .21 | .27 | .30 | .33 | .25 | -.22 | -.16 |
| | .27 | .33 | -.01 | .07 | .10 | .09 | .12 | -.13 | -.09 | -.08 | -.16 | -.08 | -.11 | .21 |
| ANI (PACU) & PSI | .13 | -.12 | .27 | -.44 | -.42 | -.50 | -.64 | -.31 | -.05 | -.04 | .08 | .06 | .11 | .19 |
| | -.11 | -.38 | .17 | .25 | .16 | .21 | .19 | .11 | .13 | .04 | -.06 | -.16 | .10 | -.16 |
| ANI (PACU) & Time to Aldrete | -- | -- | -- | -- | -- | -- | -- | -- | -- | -- | -- | -- | .32 | |
| | -- | -- | -- | -- | -- | -- | -- | -- | -- | -- | -- | -- | -.20 | |

D. ANI (PACU) Benjamini-Hochberg (FDR) corrected *P*-values (Pfol N=17; Rem N=14)

| Measures / States | Baseline EC | Baseline EO | After Loss of consciousness LOC1 | LOC2 | LOC3 | LOC4 | LOC5 | Before Recovery of consciousness ROC1 | ROC2 | ROC3 | ROC4 | ROC5 | Emergence EC | Emergence EO |
|---|---|---|---|---|---|---|---|---|---|---|---|---|---|---|
| ANI (PACU) & PLE (δ) | 0.382 | 0.829 | 0.848 | 0.280 | 0.168 | 0.168 | 0.168 | 0.382 | 0.437 | 0.280 | 0.382 | 0.382 | 0.235 | 0.829 |
| | 0.777 | 0.702 | 0.738 | 0.738 | 0.738 | 0.738 | 0.738 | 0.702 | 0.702 | 0.702 | 0.702 | 0.702 | 0.702 | 0.702 |
| ANI (PACU) & PLE (θ) | 0.697 | 0.959 | 0.301 | 0.959 | 0.959 | 0.959 | 0.959 | 0.697 | 0.697 | 0.301 | 0.569 | 0.697 | 0.697 | 0.697 |
| | 0.888 | 0.888 | 0.888 | 0.888 | 0.888 | 0.888 | 0.888 | 0.888 | 0.888 | 0.888 | 0.888 | 0.888 | 0.888 | 0.888 |
| ANI (PACU) & PLE (α) | 0.848 | 0.814 | 0.784 | 0.814 | 0.814 | 0.943 | 0.848 | 0.814 | 0.814 | 0.831 | 0.807 | 0.807 | 0.848 | 0.943 |
| | 0.661 | 0.661 | 0.661 | 0.661 | 0.661 | 0.661 | 0.661 | 0.661 | 0.661 | 0.982 | 0.879 | 0.830 | 0.661 | 0.661 |
| ANI (PACU) & PLE (β) | 0.783 | 0.820 | 0.783 | 0.783 | 0.783 | 0.977 | 0.783 | 0.783 | 0.783 | 0.783 | 0.977 | 0.783 | 0.783 | 0.783 |
| | 0.947 | 0.947 | 0.947 | 0.947 | 0.947 | 0.974 | 0.947 | 0.974 | 0.974 | 0.974 | 0.974 | 0.947 | 0.974 | 0.947 |
| ANI (PACU) & PLI (δ) | 0.778 | 0.300 | 0.311 | 0.284 | 0.284 | 0.154 | 0.284 | 0.284 | 0.154 | 0.154 | 0.341 | 0.430 | 0.154 | 0.300 |
| | 0.942 | 0.942 | 0.942 | 0.942 | 0.942 | 0.962 | 0.942 | 0.942 | 0.942 | 0.942 | 0.962 | 0.942 | 0.942 | 0.962 |
| ANI (PACU) & PLI (θ) | 0.910 | 0.910 | 0.396 | 0.680 | 0.910 | 0.910 | 0.910 | 0.443 | 0.910 | 0.205 | 0.205 | 0.140 | 0.443 | 0.910 |
| | 0.992 | 0.992 | 0.992 | 0.992 | 0.014 | 0.992 | 0.992 | 0.378 | 0.992 | 0.992 | 0.992 | 0.992 | 0.992 | 0.992 |
| ANI (PACU) & PLI (α) | 0.983 | 0.513 | 0.513 | 0.513 | 0.859 | 0.927 | 0.685 | 0.513 | 0.513 | 0.755 | 0.513 | 0.513 | 0.513 | 0.513 |
| | 0.892 | 0.733 | 0.733 | 0.733 | 0.733 | 0.733 | 0.892 | 0.892 | 0.892 | 0.892 | 0.733 | 0.733 | 0.733 | 0.892 |
| ANI (PACU) & PLI (β) | 0.727 | 0.601 | 0.562 | 0.364 | 0.275 | 0.371 | 0.896 | 0.727 | 0.896 | 0.601 | 0.601 | 0.371 | 0.259 | 0.259 |
| | 0.881 | 0.881 | 0.881 | 0.881 | 0.881 | 0.881 | 0.917 | 0.881 | 0.881 | 0.881 | 0.917 | 0.881 | 0.881 | 0.881 |
| ANI (PACU) & power (δ) | 0.828 | 0.927 | 0.927 | 0.483 | 0.042 | 0.028 | 0.028 | 0.616 | 0.548 | 0.548 | 0.548 | 0.548 | 0.828 | 0.828 |
| | 0.965 | 0.965 | 0.965 | 0.965 | 0.965 | 0.965 | 0.965 | 0.965 | 0.965 | 0.965 | 0.965 | 0.616 | 0.965 | 0.965 |
| ANI (PACU) & power (θ) | 0.658 | 0.952 | 0.952 | 0.952 | 0.429 | 0.429 | 0.429 | 0.952 | 0.952 | 0.952 | 0.952 | 0.952 | 0.952 | 0.952 |
| | 0.948 | 0.684 | 0.684 | 0.804 | 0.684 | 0.804 | 0.684 | 0.684 | 0.684 | 0.684 | 0.684 | 0.830 | 0.684 | 0.804 |
| ANI (PACU) & power (α) | 0.828 | 0.828 | 0.828 | 0.828 | 0.644 | 0.658 | 0.658 | 0.828 | 0.828 | 0.828 | 0.828 | 0.828 | 0.828 | 0.828 |
| | 0.797 | 0.797 | 0.797 | 0.797 | 0.797 | 0.797 | 0.797 | 0.797 | 0.797 | 0.830 | 0.797 | 0.950 | 0.797 | 0.797 |
| ANI (PACU) & power (β) | 0.768 | 0.559 | 0.427 | 0.523 | 0.427 | 0.427 | 0.427 | 0.531 | 0.523 | 0.523 | 0.523 | 0.531 | 0.531 | 0.577 |
| | 0.881 | 0.881 | 0.982 | 0.881 | 0.881 | 0.881 | 0.881 | 0.881 | 0.881 | 0.881 | 0.881 | 0.881 | 0.881 | 0.881 |
| ANI (PACU) & PSI | 0.877 | 0.877 | 0.709 | 0.319 | 0.319 | 0.301 | 0.070 | 0.641 | 0.877 | 0.877 | 0.877 | 0.877 | 0.877 | 0.877 |
| | 0.848 | 0.848 | 0.848 | 0.848 | 0.848 | 0.848 | 0.848 | 0.848 | 0.848 | 0.892 | 0.892 | 0.848 | 0.848 | 0.848 |
| ANI (PACU) & Time to Aldrete | -- | -- | -- | -- | -- | -- | -- | -- | -- | -- | -- | -- | 0.541 | |
| | -- | -- | -- | -- | -- | -- | -- | -- | -- | -- | -- | -- | 0.547 | |

**Table S3.** Pearson's correlation between ANI and results values.

Pearson's correlation coefficients and Benjamini-Hochberg (FDR) adjusted *P*-values of corresponding coefficients, whereby **(A)** and **(B)** include correlation between ANI values and results values (connectivity, powers at four frequency bands, PSI, and time to Aldrete 9), **(C)** and **(D)** include correlation between ANI at PACU (averaged values of EC and EO emergence) and results values. The blue and red values represent the correlation values in propofol and remimazolam groups, respectively. The highlights colored with 2-steps correspond to the significance level of the FDR-corrected *P*-value (yellow: $P < 0.05$, orange: $P < 0.01$).

| Score | Term | Description |
|---|---|---|
| +4 | Combative | Overtly combative, violent, immediate danger to staff |
| +3 | Very agitated | Pulls or removes tube(s) or catheter(s) |
| +2 | Agitated | Frequent non-purposeful movement, fights ventilator |
| +1 | Restless | Anxious but movements not aggressive vigorous |
| 0 | Alert and calm | |
| -1 | Drowsy | Not fully alert, but has sustained awakening (eye-opening/eye contact) to voice ( ≥10 seconds) |
| -2 | Light sedation | Briefly awakens with eye contact to voice ( <10 seconds) |
| -3 | Moderate sedation | Movement or eye opening to voice (but no eye contact) |
| -4 | Deep sedation | No response to voice, but movement or eye opening to physical stimulation |
| -5 | Unarousable | No response to voice or physical stimulation |

**Table S4.** Richmond Agitation Sedation Scale (RASS)

RASS scores evaluate alertness. 0 = alert, negative value = drowsy.

| Literature | Monitoring System (range) | Dose at induction (number of patients) | R vs. P during unconscious (before extubation) | R vs. P during emergence (POD0) | R vs. P at POD1 |
|---|---|---|---|---|---|
| Doi and colleagues (2020)[5] | BIS<br>R-6mg (40-82)<br>R-12mg (47.8-84)<br>P (39-56.3) | R (150): 6mg/kg/hr<br>R (150): 12mg/kg/hr<br>(flumazenil given if not awaken)<br>P (75): 2.0-2.5 mg/kg | R > P (sig.) | P > R (6mg and 12 mg) in BIS. Difference between 20 - 40min after the end of IMP. | n.a. |
| Choi and colleagues (2022)[6] | PSI (25-50) | R (70): 6mg/kg/hr;<br>P (69): 5mcg | R > P (sig.) | n.a. (immediately after extubation – no group difference) | No group difference on quality of recovery |
| Shi and colleagues (2022)[7] | BIS (45-60) | R (38): 0.2mg/kg (flumazenil given)<br>P (38): 2mg/kg | R > P (sig.) | (1 minute after extubation) R > P<br>Time to extubation and time at PACU shorter in R | n.a. |
| Zhang and colleagues (2023)[8] | BIS (40-60) | R (71): 0.1mg/kg/hr;<br>P (71):1-1.5mg/kg | n.a. | Not monitored in BIS, but there was no group difference on time to emergence. | No group difference |
| Tang and colleagues (2023)[9] | BIS (50-60) | R (56): 6mg/kg/hr;<br>P (58): 3.5mcg | R > P (sig.) | Not monitored in BIS, but other indices showed slower recovery in R. | Significantly higher quality of recovery in R |
| Kim and colleagues (2024)[10] | BIS (unknown; 50-60) | R (26): 6mg/kg/hr; (flumazenil given)<br>P (26): 3.5mcg | n.a. | (at least 30 min after end of surgery); no group difference. | n.a. |
| Present study | PSI (40) | R (24): 6mg/kg/hr;<br>P (24): 4mcg | R > P (sig.) | (immediately during emergence) P > R. Time to Aldrete shorter in P. | n.a. |

**Table S5.** Summary of relevant literature comparing between Remimazolam- and Propofol-induced anaesthesia.

Comparison includes depth of anesthesia, speed of emergence, and quality of recovery between the two anaesthetic agents at various periods. R: Remimazolam, P: Propofol. Inequality symbols (< or >) represent whether values from monitoring system (BIS or PSI) are greater/smaller than the other.

| | Propofol | Remimazolam |
|---|---|---|
| | Mean MAE | |
| RNF | 7.714286 | 4.885882353 |
| Linear | 5.104974 | 5.034812725 |

**Table S6.** Mean absolute error (MAE) for both groups.

RNF: Random forest regression, Linear: Linear regression.
